# *Ex vivo* drug testing in an ultra-rare sarcoma reveals therapeutic vulnerability and resistance

**DOI:** 10.1101/2022.08.03.22278327

**Authors:** Sharon Pei Yi Chan, Baiwen Luo, Benjamin Jieming Chen, Andre Villanueva, Sam Xin Xiu, Benjamin Livingstone Farah, Nicholas Shannon, Chin-Ann Johnny Ong, Claramae Shulyn Chia, Ming-Hui Yong, Krishan Kumar, London Lucien Ooi, Timothy Kwang Yong Tay, Xing Yi Woo, Tan Boon Toh, Edward Kai-Hua Chow, Valerie Shiwen Yang

## Abstract

Solitary fibrous tumors (SFTs) are rare soft tissue sarcomas for which therapeutic options are limited and ineffective. We successfully demonstrated how functional personalized treatment was implemented in the clinic for an ultra-rare sarcoma with otherwise limited options, through a combined strategy of patient-derived model development and computational drug analytics. Molecular profiling of tumours and patient-derived models uncovered potential biomarkers to predict responses to specific drugs.

We generated patient-derived SFT cells (PDSC) and used a computational combinatorial drug screening analytics platform, Quadratic Phenotypic Optimization Platform (QPOP), to determine therapeutic vulnerability and resistance in an ultra-rare locally recurrent brain SFT and its distant liver metastasis. QPOP derived and ranked the efficacy of 531,441 drug combinations, revealing BETi-pazopanib synergy in the liver lesion that outperforms standard-of-care combination doxorubicin-ifosfamide, which was antagonistic. In tumour and PDSC from the pazopanib-resistant brain lesion, transcriptomic analyses identified the UGT1A family as potential biomarkers of pazopanib resistance. Eribulin sensitivity was predicted to be shared across both lesions. Our patient was therefore treated with eribulin and successfully gained clinically meaningful disease control.

## BACKGROUND

Solitary fibrous tumor (SFT) is a rare soft tissue sarcoma (STS) representing four percent of all STS and less than one percent of all cancers (1,2). They comprise a histologic spectrum of fibroblastic mesenchymal neoplasms often presenting as a slow growing mass in adult patients (3). Though prototypically described to be originating from the visceral pleura, it can ubiquitously affect any anatomic site (4). The diagnosis of SFT is established by characteristic histologic and immunohistochemical features: STAT6 nuclear expression, diffuse and strong reactivity to CD34, and supported by positive findings for CD99 and BCL-2 (5,6). Genetically, SFTs harbour recurrent intrachromosomal 12q13 rearrangements, leading to an in-frame fusion of the genes *NAB2* (NGFI-A-Binding protein 2) and *STAT6* (Signal Transducer and Activator of Transcription 6) (7,8). This fusion drives constitutional upregulation of oncogenic *EGR1* (Early Growth Response 1)-dependent gene expression leading to a cascade of events causing nuclear differentiation and proliferation (9). The intragenic breakpoints within the *NAB2* and *STAT6* gene loci are variable, with more than 40 breakpoint variants reported and associated with distinct clinical features (10). The most common and canonical *NAB2* exon 4-*STAT6* exon 2 fusion variant corresponds to classic pleuropulmonary SFTs with mostly benign behavior and occurs in older patients (11). In contrast, *NAB2* exon 6-*STAT6* exon 16/17, the second most common fusion variants, represents typical “hemangiopericytoma” with an aggressive phenotype (11).

Localized SFTs with classical morphologic features are usually clinically indolent with favourable prognoses. However, the behaviour of SFT is unpredictable and can progress to aggressive neoplasms characteristic of high-grade sarcomas (12,13). Like many other soft tissue tumors, surgical management is the mainstay for SFT. Despite local control following complete surgical excision with good margins, recurrence can occur in up to 10-40% of SFT, with metastatic spread to lungs (31%), liver (24%), and bone (15%)(12,13). For this subset of patients with unresectable or metastatic disease, options for effective treatment are particularly limited. Radiation therapy and chemotherapy regimens have not demonstrated global effectiveness, and thus no standardized treatments have been identified. Anthracycline-based regimens for advanced or metastatic STS involving treatment with doxorubicin as monotherapy or in combination with other cytotoxic drugs (e.g. ifosfamide) have been advocated as first-line chemotherapy (13), with a retrospective cohort reporting progression free survival of three to five months in line with other STSs (14). Given that SFTs are highly vascularized tumors with high expression rates of angiogenic markers, the efficacy of anti-angiogenic agents have been explored in SFT with retrospective and prospective evidence suggesting efficacy for unresectable disease (15,16). Temozolomide-bevacizumab combination therapy (17,18) as well as sunitinib (19) have produced tumor response and disease control in some patients with advanced SFTs. Sorafenib (20) and pazopanib (21–23) have also showed promising response rates compared to those observed for standard chemotherapy. Given the rarity of SFTs, preclinical or clinical data is restricted to case reports or small retrospective cohorts, with few randomized trials. High quality evidence is lacking and there are still no consensus clinical guidelines to guide clinicians in treating this aggressive, ultra-rare disease (15,23,24).

Rational design of drug combinations has become a promising approach to evaluate drug sensitivity and resistance in cancer treatment. We had previously developed a mechanism-agnostic, quantitative, phenotype-based drug screening method – Quadratic Phenotypic Optimization Platform (QPOP) that was successfully applied in bortezomib-resistant multiple myeloma (25,26) and T-cell lymphoma (27). QPOP is a rational, experimental data-driven platform that designs effective combination therapy for a specific disease model without prior reference to mechanistic assumptions. By utilizing small experimental datasets to interrogate the entire drug-dose search space, QPOP circumvents the need for exhaustive testing to efficiently identify the best drug combination (28). An orthogonal array composite design (OACD) computes minimum test combinations to be used as experimental data points for subsequent model fitting and drug screening (28). Patient sample responses to combinations of drugs that fit within the OACD can be used to derive patient-specific coefficients, thereby serving as a predictor of tumor response to all potential drug combinations. In this aggressive SFT that has failed multiple lines of treatment with uncertain therapeutic options, we employed the use of QPOP to systematically evaluate drug combinations in matched SFT samples and provide a rational approach to address this lack of effective therapeutic options. Our study demonstrates proof-of-concept for the QPOP platform to identify drug combinations, and single drugs, against an ultra-rare sarcoma that is otherwise resistant to standard of care, supporting the utility of phenotypic-driven drug discovery for personalized medicine in this setting.

## METHODS

### Patient tissue and clinical data collection

Clinical information and imaging was retrieved from electronic medical records. All histological parameters were reviewed by sarcoma pathologists. The diagnosis was rendered according to the World Health Organization (WHO) classification (29) and updated with reference to WHO 2016 criteria (30). Written informed consent was obtained from the patient for use of biospecimens and clinical data was obtained in accordance with the Declaration of Helsinki. Tissue collection and consent protocols were governed by ethics approval from the SingHealth Centralized Institution Review Board (CIRB-2018/3182). QPOP analysis of patient samples was performed in accordance with ethics approval from NUS Institutional Review Board (H-18-032).

### Isolation, culture and characterization of patient-derived sarcoma cells

Freshly excised tumor samples were digested with a GentleMACS dissociator in warm RPMI-1640 medium (Biowest) with 100 ug/mL LiberaseTM (Sigma Aldrich, Singapore) for five minutes and then incubated on a shaker at 70 rpm in a cell culture incubator for two hours. Primary patient-derived sarcoma cells (PDSCs) were collected after filtration and centrifugation. Isolated PDSCs were cultured in RPMI-1640 medium supplemented with 20% (v/v) fetal bovine serum (FBS; Gibco, USA), 1% (v/v) penicillin/streptomycin (Gibco, USA). Proliferation profiles of PDSCs were determined using alamarBlue (AB) assay (Life Technologies, Singapore) at timepoints of day one, three, five and seven. Cell numbers were calculated by seeding known quantities of cells and correlation with fluorescence emission. *In vitro* cell viability was performed using the LIVE/DEAD viability/cytotoxicity assay (Life Technologies, Singapore). Briefly, PDSCs were stained in LIVE/DEAD working solution (2 µM Calcein AM and 4 µM ethidium homodimer-1 dye in DPBS) for 20 minutes inside a cell culture incubator and visualized using a fluorescence microscope (EVOS M7000, Life Technologies, Singapore). To verify the successful isolation and culture of SFT cells, immunofluorescence staining of key SFT markers was performed. Cells were washed with phosphate buffered saline (PBS), fixed using 4% paraformaldehyde in PBS for 15 minutes, and permeabilised with 0.1% Triton X-100 in PBS for two minutes. Blocking was performed with 2.5% FBS in PBS for 1 hour at room temperature. Primary antibodies were diluted in the blocking reagent and incubated with the samples overnight at 4°C. Following primary antibody incubation, samples were washed in PBS and incubated with secondary antibody for one hour at room temperature. The following primary antibodies and dilutions were used: Vimentin (Abcam, 1:400), BCL2 (Abcam, 1:200), CD34 (Abcam, 1:200), ALDH1 (Abcam, 1:200), and STAT6 (Abcam, 1:200), while secondary antibodies used were goat anti-mouse IgG H&L (Alexa Fluor® 488; 1:500) and goat anti-rabbit IgG H&L (Alexa Fluor® 594, 1:500). PDSCs were then mounted with a DAPI-contained mounting medium (Abcam) on a glass slide and visualized under a fluorescence microscope (EVOS M7000, Life Technologies, Singapore).

### *Ex vivo* drug testing and analytics

A QPOP-based protocol was used to identify optimal clinically-applicable combinatorial drug regimens. Pre-experimentally selected drug candidates are shown in Table 1. A 12-drug, three-dose level QPOP analysis consisting of 155 experimental combinations (Table S1) was performed, where the three levels corresponded to concentrations of the drugs at their IC_0_, IC_10_ and IC_20_ derived from the mean of a panel of five STS cell lines (Table 1).

**Table 1:** List of drugs, their targets and doses used for QPOP analysis on patient-derived sarcoma cells.

|  | Drug | Target | IC<sub>0</sub> (μM) | IC<sub>10</sub> (μM) | IC<sub>20</sub> (μM) | C<sub>max</sub> (μM)* |
| --- | --- | --- | --- | --- | --- | --- |
| D1 | Ifosfamide | DNA (alkylation) | 0 | 2.083 | 4.167 | 431 |
| D2 | Doxorubicin | Topoisomerase II | 0 | 0.0516 | 0.1032 | 6.73 |
| D3 | Panobinostat | HDAC | 0 | 0.0041 | 0.0082 | 0.082 |
| D4 | Gemcitabine | DNA | 0 | 0.00725 | 0.0145 | 89.3 |
| D5 | 5-Azacytidine | DNMT | 0 | 0.1535 | 0.307 | 3.07 |
| D6 | Roybinostat | HDAC | 0 | 0.852 | 1.704 | - |
| D7 | OTX015 | BET | 0 | 2.126 | 4.25 | - |
| D8 | Palbociclib | CDK4/6 | 0 | 0.00505 | 0.0101 | 0.101 |
| D9 | Pazopanib | Multityrosine kinases | 0 | 6.65 | 13.3 | 133 |
| D10 | Eribulin mesylate | Microtubule | 0 | 0.0137 | 0.0273 | 0.508 |
| D11 | Trabectedin | DNA | 0 | 0.00012 | 0.00024 | 0.0024 |
| D12 | Ponatinib | Multityrosine kinases | 0 | 0.00685 | 0.0137 | 0.137 |

Early passage PDSCs (passage two) were plated in 384-well plates (Corning) before undergoing either a QPOP-specific drug combinatorial treatment or serial dose response assays. These combinations were prepared with the aid of a liquid handler (Mini Janus, PerkinElmer) or a digital dispenser (D300e, Tecan). After 48-hour drug treatment, CellTiter-Glo® Luminescent Cell Viability Assay was used to quantify the cell viability, and the normalized viability scores were used as inputs for QPOP second-order regression analysis as described by Rashid et al. (2018) (25). The second-order quadratic equation is as follows:

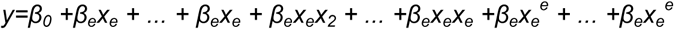

where *y* represents the desired output, *x*_*e*_ is the nth drug dosage, *β*_*e*_ is the intercept term, *β*_*e*_ is the single-drug coefficient of the *e*^*e*^ drug, *β*_*e*_ is the interaction coefficient between the *e*^*e*^ and *e*^*e*^ drugs, and *β*_*e*_ is the quadratic coefficient for the *e*^*e*^ drug. Based on these experimental data points, QPOP analysis generated all possible permutations of these combinations and their corresponding projected output values, as well as parabolic response surface maps depicting interactions between two drugs on cell viability. All two-drug and three-drug combinations were compared and ranked in order of efficacy.

### Validation of QPOP-ranked efficacious drug combinations

#### Determination of Combination Index

Single-drug and combination dose-response assays were conducted to determine the IC_50_ values of each drug as monotherapy and in combination. For the combination dose-response assay, both drugs were administered together in a fixed dose ratio across the range of concentrations tested. To compute IC_50_ values, sigmoidal dose-response curves were generated by fitting calculated cell viability values at different log concentrations using GraphPad Prism (GraphPad Software). With reference to the CI theorem of Chou-Talalay, the CI values were subsequently computed from the dose-response curves and IC_50_ values (31). The resulting CI score offers quantitative definition for additive effect (CI = 1), synergism (CI < 1), and antagonism (CI > 1) in drug combinations.

#### Immunofluorescence assay and quantification

PDSCs were seeded in 96-well plates at a density of 500 cells per well. Cells were allowed to adhere and recover for 24 hours prior to drug treatment. Thereafter, cells were treated with 10μM of BET inhibitor OTX015 and/or 5μM Pazopanib as single agents or in combination and incubated for 48 hours. Cells were then prepared for immunofluorescence assay as described above. Primary antibodies against C-MYC (Abcam, 1:100) and C-KIT (Abcam, 1:400) and Alexa Fluor 488-conjugated secondary antibodies (1:200) were used. Samples were counterstained with 4’,6-diamidino-2-phenylindole (DAPI) and visualized by a confocal laser scanning microscope (LSM 880 Airy Scan, Carl Zeiss). In determining the level of cellular fluorescence, the following formula was used to calculate the corrected total cell fluorescence (CTCF): CTCF = Integrated Density – (Area of selected cell X Mean fluorescence of background readings).

### RNA isolation and whole transcriptome sequencing

Total RNA was extracted from PDSCs untreated, treated singly or combination. PDSCs were seeded, allowed to adhere and recover for 24 hours prior to drug treatment. Cells were incubated for 16 hours with the above drugs at the stated concentrations. The 16-hour timepoint was chosen to minimise cell cycle arrest targets in analysis and facilitate detection of early response markers. Total RNA was extracted and purified with RNeasy Mini kit (Qiagen). RNA concentration was determined by Nanodrop 1000 Spectrophotometer (ThermoFisher Scientific). Extracted RNA was converted into RNA sequencing library using the NEBNext® Ultra™ Directional RNA Library Prep Kit (NEB) using manufacturer’s instructions. Whole transcriptome sequencing of cell lines was performed on the Illumina NovaSeq platform (Novogene, Singapore) using the standard Illumina RNA-seq protocol. Paired-end raw sequencing reads were trimmed with Trim Galore (version 0.4.2_dev; https://www.bioinformatics.babraham.ac.uk/projects/trim_galore/) with the following parameters: -trim-n -paired (paired-end). Cleaned reads were then mapped to the human hg38 using the RSEM pipeline (version 1.1.11). Read counts were obtained for each sample and edgeR (version 3.34.1) was used for differential gene expression analyses with default settings. Only genes that were expressed (Count Per Million ≥ 1) in at least two samples were included for analysis. Differential expression was calculated using the exactTest function with dispersion = 0.04. Untreated and DMSO treated samples were grouped as biological replicates. Genes are differentially expressed if they show at least a twofold change in expression with a false discovery rate (FDR) < 0.05 after correcting for multiple testing by the Benjamini-Hochberg method (32).

### *NAB2-STAT6* fusion gene analysis

STAR-Fusion (1.10.1) integrated into the STAR aligner (2.7.8a) running on the provided Singularity (3.9) container was used to identify potential fusion genes. STAR-Fusion takes in trimmed, paired end RNAseq reads and aligns the reads using the STAR aligner, refining reads that span or straddle fusion junctions. Gene fusion candidates were filtered to retain fusions with at least five fusion junction reads. Reads deposited in the chimeric-reads file indicate putative fusions detected.

### Statistical analysis

All experiments were performed in at least duplicate technical repeats, with data presented as means ± standard deviation (SD). Student’s two-tailed t test was used for the comparison of two independent groups. A *p* value of <0.05 was accepted as statistically significant. GraphPad Prism 8 software was used for data analysis.

## RESULTS

### Clinical presentation and pathological findings

A young female in her 20s with an unremarkable past medical history presented to an external hospital with severe headaches. Magnetic Resonance Imaging (MRI) of the brain showed a left lateral ventricular mass with midline shift. Histological evaluation upon excision of the mass revealed a WHO Grade II HPC/ SFT. Within the next 6 years, the patient developed multiple local recurrences and metastatic disease to bone and liver (Fig. 1A, 1B, S1). Her diagnosis was revised to a higher grade HPC/SFT (Grade III) with elevated mitoses at 5 per 10 HPFs, invasion of the cerebral cortex, minimal intervening collagenous stroma between the tumor fascicles, geographic necroses and staghorn vessels (Fig. 1B). She was given two cycles of liposomal doxorubicin. Repeat imaging showed disease progression in her liver though brain lesions remained stable (Fig. S2). She subsequently underwent a palliative resection of her dominant liver metastasis and commenced palliative pazopanib as second-line systemic treatment. Repeat imaging two months later showed mixed response in her liver lesions and progression in her dominant brain lesion which remained asymptomatic (Fig. S2). She then underwent yttrium-90 radioembolization to consolidate local control of her liver lesions, as well as palliative radiotherapy to her bone lesions, and continued pazopanib. Four months later, given the isolated site of progression in the brain with disease control elsewhere, she underwent her third craniotomy with palliative intent and was resumed on pazopanib post-operatively. Repeat imaging showed continued response in her liver lesions and stable disease in her bone lesions, though MRI of the brain revealed a small enhancing nodule suspicious for residual lesion (Fig. S2). She remains clinically well at present. Oncomine Comprehensive Panel v3 (RNA) and Oncomine Comprehensive Assay Plus (DNA) assays performed on her primary brain lesion as well as liver metastasis did not reveal any clinically actionable findings (Fig. S3), supporting the need for an alternative strategy to identify effective drugs in her multiply resistant tumors. A portion of her dominant liver metastasis and locally recurrent brain lesion, resected in April 2021 and December 2021 respectively, were dissociated in our laboratory and used for *ex vivo* drug testing and generation of early passage patient-derived cell lines. The dominant liver metastasis and locally recurrent brain lesion are referred to as hSC06 and hSC31 respectively (Fig. 1C).

**Figure 1.**
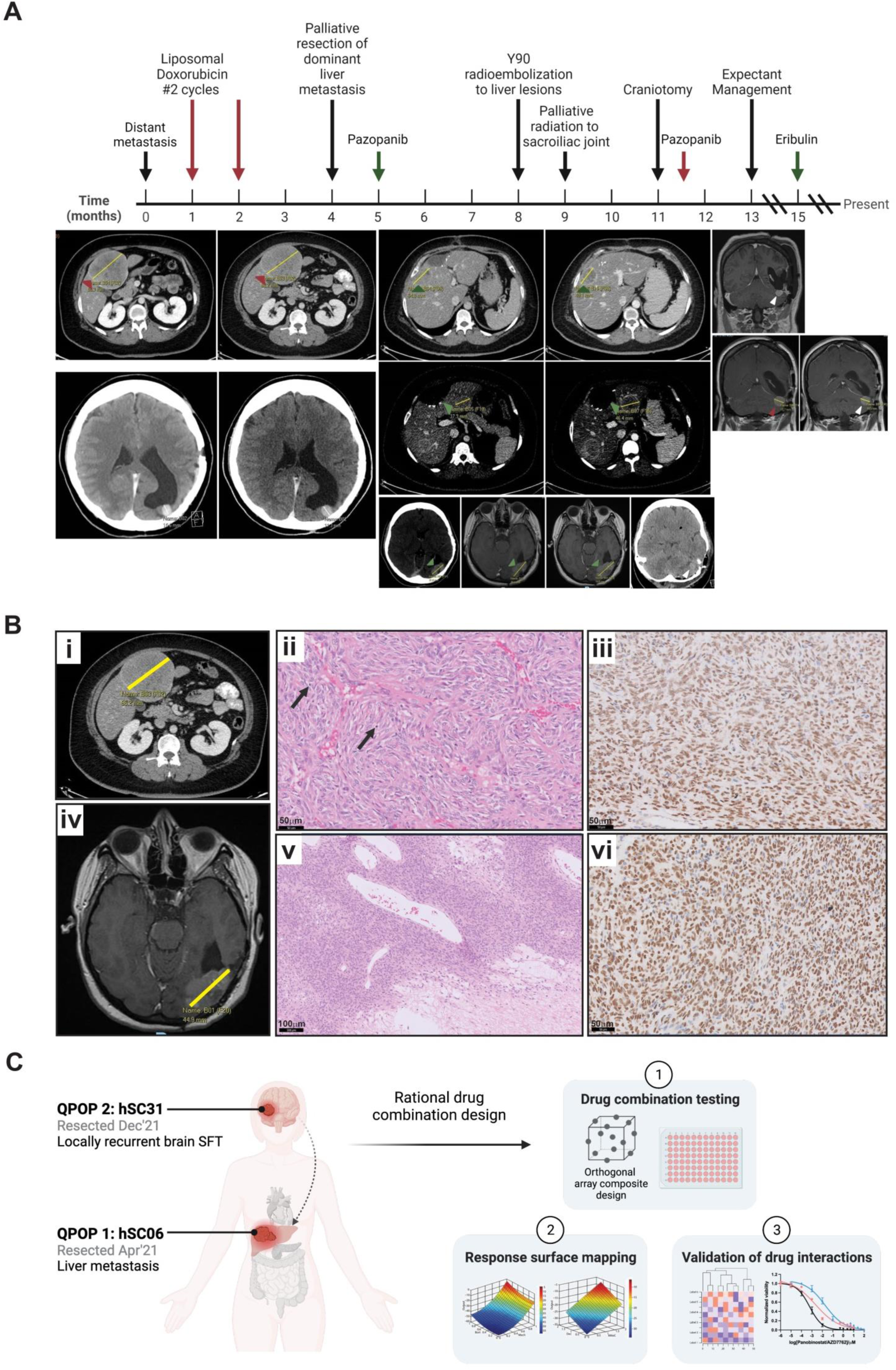
Clinical case presentation and overview of the QPOP process and analytics. **(A)** Clinical course of our patient presenting with rare and atypical malignant solitary fibrous tumor (SFT) who failed first-line chemotherapy. **(B) (i)** Computed tomography (CT) imaging of the dominant liver metastasis. **(ii)** Haematoxylin and eosin (H&E) stained section of the liver metastasis demonstrating a tumor comprised of cellular sheets of spindled tumor cells arranged in ill-defined fascicles, with only small amounts of collagen, as well as “staghorn” vessels arising from the liver, with increased mitotic figures, marked with arrows (200x magnification, 20x objective), and **(iii)** STAT6 immunoperoxidase stained section of the liver metastasis showing diffuse nuclear immunoreactivity (200x magnification, 20x objective). **(iv)** Magnetic resonance imaging (MRI) of the locally recurrent primary brain lesion. **(v)** Haematoxylin and eosin stained section of the re-excised brain SFT demonstrating markedly cellular sheets of spindled tumor cells arranged in ill-defined fascicles, with only small amounts of collagen, as well as dilated vessels and areas of necrosis (100x magnification, 10x objective), and **(vi)** STAT6 immunoperoxidase stained section of the brain mass showing diffuse nuclear reactivity (100x magnification, 10x objective). **(C)** The dominant liver metastasis (hSC06) was first resected and screened by QPOP (QPOP 1; April 2021), followed by the locally recurrent brain SFT (hSC31) from a palliative resection of the occipital region (QPOP 2; December 2021). The QPOP workflow involves 1) experimental testing of 155 drug combinations generated by orthogonal array composite design (OACD) from 12 drugs at three concentration levels; 2) measurement of hSC06 and hSC31 cell viability at 48 hours post-treatment to be used as phenotypic output for fitting a second order linear regression; and 3) validation studies on QPOP-identified top- and bottom-ranking drug combinations.

### Establishment and characterisation of patient-derived SFT cell lines

Tumor samples were processed immediately upon collection. Morphologically, cells were elongated and spindle-shaped, growing in loose colonies (Fig 2A). Cell viability assay revealed negligible amounts of dead cells in both lines when maintained in culture for seven days (Fig. 2A, B). Both hSC06 and hSC31 demonstrated proliferative capacity over seven days with a doubling time of five to seven days (Fig. 2B). Ki67 also indicated the presence of cycling and mitotically active cells (Fig. 2C). To establish if SFT characteristics of the locally recurrent brain lesion and liver metastasis were retained in our PDSCs, we looked for molecular markers by immunostaining. Both hSC06 and hSC31 were positive for key SFT markers Vimentin, BCL2, CD34, ALDH1, and STAT6 (Fig. 2C). The liver metastasis hSC06 was strongly positive for Vimentin, a key epithelial-mesenchymal transition (EMT) marker, reflective of its metastatic potential. STAT6, a surrogate for the detection of the hallmark *NAB2-STAT6* gene fusion (5,33,34), showed diffuse nuclear positivity in both hSC06 and hSC31 (Fig. 2C). CD34-negative SFTs have been associated with metastatic disease (35) and a higher proportion of CD34-positive cells in the locally recurrent brain lesion (hSC31) compared to the liver metastasis (hSC06, Fig. 2C) supports this finding. Diffuse cytoplasmic staining of BCL2 in both lines is also highly characteristic of SFT (36). ALDH1, a stem cell marker, is used to distinguish SFT from other histologically similar tumors such as meningiomas and synovial sarcomas (37). Cytoplasmic ALDH1 staining in both hSC06 and hSC31 further supports the retention of SFT characteristics in our PDSCs.

**Figure 2.**
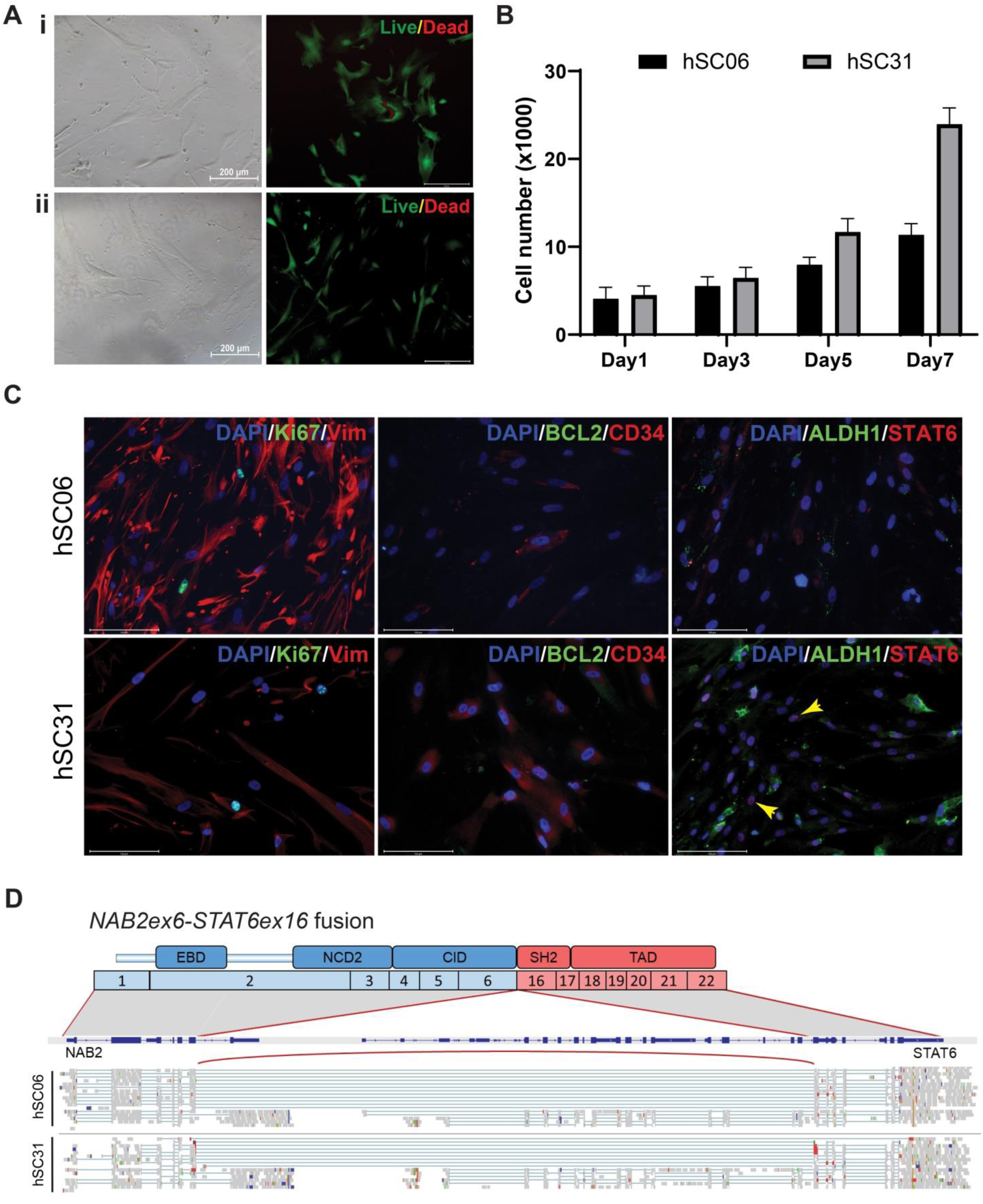
Establishment and characterisation of patient-derived sarcoma cells (PDSCs). **(A)** Representative phase-contrast micrographs of PDSCs from **(i)** distant liver metastasis (hSC06) and **(ii)** locally recurrent brain SFT (hSC31) demonstrate long spindle-shaped cells growing in loose colonies. LIVE/DEAD imaging of PDSCs after seven days is shown. Green: live cells; red: dead cells. **(B)** alamarBlue™ cell proliferation assay demonstrates proliferative capacity of PDSCs over seven days in culture. **(C)** Immunofluorescence staining for key SFT markers Vimentin, BCL2, CD34, ALDH1, and STAT6. Scale bar = 150 μm. **(D)** Schematic of the NAB2-STAT6 gene fusion detected in hSC06 and hSC31 by paired-end transcriptome sequencing.

Detection of the *NAB2-STAT6* fusion gene is the gold standard for SFT diagnosis. The *NAB2-STAT6* gene fusion was indeed detected from transcriptomic analysis of both hSC06 and hSC31, confirming that both patient-derived lines were indeed SFT. Analysis of intragenic breakpoints within the *NAB2* and *STAT6* gene loci revealed fusion of *NAB2* exon 6 to *STAT6* exon 16 in both hSC06 and hSC31, resulting in the expression of the fusion gene *NAB2* exon 6-*STAT6* exon 16 (6-16) (Fig. 2D). Consequently, the C-terminal Interacting Domain (CID) of NAB2, which plays an inhibitory role, is truncated and fused with the Trans-Activation Domain (TAD) of STAT6, leading to a fusion product that drives the overexpression of *EGR1*. The 6-16 variant has been reported to occur more often in extra-thoracic regions and meninges, and is associated with an aggressive clinical phenotype (11), consistent with clinical course observed in our patient.

### Ex vivo QPOP drug testing identified tumor-specific responses consistent with clinicoradiological findings and drug combinations superior to standard regimens

We carried out a QPOP-based protocol on hSC06 and hSC31 to systematically screen for drug sensitivity and resistance. Top five two-drug combinations across hSC06 and hSC31 were ranked in a heatmap against standard of care regimens and agents known to be active in SFT and soft tissue sarcomas (doxorubicin, pazopanib, ifosfamide, gemcitabine, trabectedin, eribulin, palbociclib). QPOP identified differential drug response across the matched samples (Fig. 3). The top ranked two-drug combinations were the BET inhibitor OTX015 and pazopanib, followed by eribulin and pazopanib in hSC06 (Fig. 3Ai) whereas eribulin and OTX015 was top-ranked for hSC31 (Fig. 3Ai). hSC06 was more sensitive towards pazopanib-based therapies, but sensitivity considerably shifted towards eribulin in hSC31 (Fig. 3A, B). Standard of care regimens ranked low in the QPOP assay (Fig. 3A). Out of a total of 288 one-and two-drug combinations, the standard of care combination regimen of doxorubicin and ifosfamide was ranked 237^th^ and 94^th^ in hCS06 and hSC31 respectively, while doxorubicin as a single agent was ranked 213^rd^ and 170^th^ in hCS06 and hSC31 respectively (Fig. 3A, B). The anti-angiogenic and multityrosine kinase inhibitor pazopanib was ranked 19^th^ and 125^th^ for hSC06 and hSC31 respectively, demonstrating enhanced killing as single agent in hSC06 as compared to hSC31 (Fig. 3Ai, 3Bi). This recapitulated what was seen clinicoradiologically: resistance to liposomal doxorubicin, and continued response in the liver lesions but progression in the residual brain lesion with systemic pazopanib treatment (Fig. 1A, S2).

**Figure 3.**
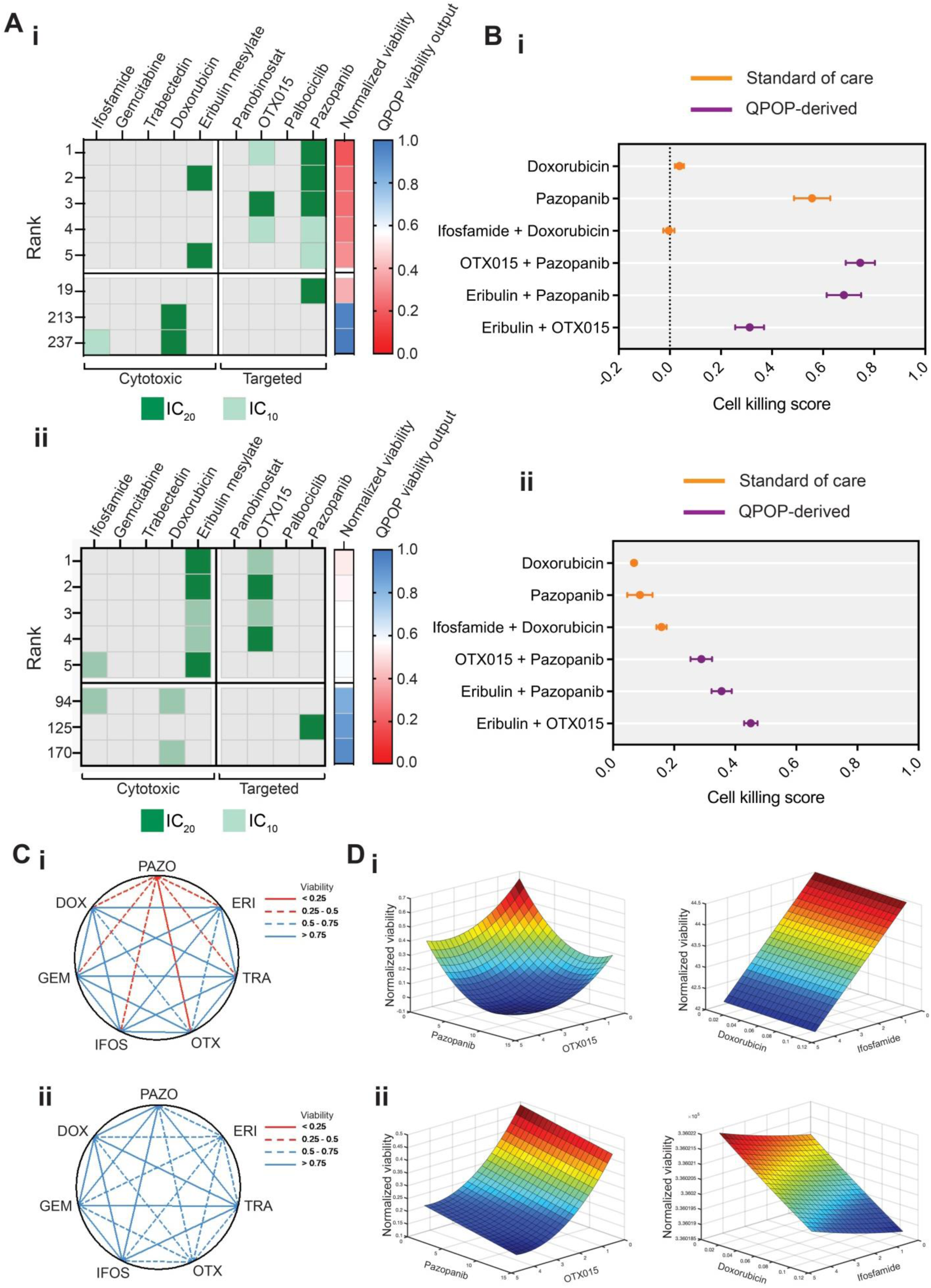
Ex vivo QPOP drug testing identified tumor-specific responses and drug combinations superior to standard regimens. **(A)** Heatmap of QPOP-derived differential drug response across **(i)** hSC06 and **(ii)** hSC31. Top five two-drug combinations were ranked according to drug efficacy against standard of care regimens in soft tissue sarcoma (doxorubicin, ifosfamide + doxorubicin, pazopanib, gemcitabine, trabectedin, eribulin, palbociclib), with their corresponding QPOP viability output (right panel). Higher concentrations used in QPOP (IC20), indicated by a darker shade of green; lower concentrations used in QPOP (IC10), indicated by a lighter shade of green. **(B)** Forest plot of cell killing scores represented between zero and one in **(i)** hSC06 and **(ii)** hSC31 comparing the relative efficacy of top-ranked 2-drug combinations against standard regimens. Standard of care regimens, orange; QPOP-derived combinations, purple. **(C)** Polygonograms showing the contrasting drug-drug interactions between **(i)** hSC06 and **(ii)** hSC31. More weighted red line is indicative of greater efficacy whereas more weighted blue line is indicative of less efficacy. **(D)** Parabolic response surface maps of **(i)** hSC06 and **(ii)** hSC31 showing drug interactions of OTX+PAZO and IFOS+DOX. Lengths of axes differ to aid visualisation.

To delineate the relative efficacy of the QPOP-derived top-ranked combinations with standard regimens, cell killing scores derived from normalized cell viability were compared (Fig. 3B). Pazopanib-based combinations showed considerably greater cell killing of hSC06 (Fig. 3Bi) compared to hSC31 (Fig. 3Bii), with more than 50% killing alone as a single agent. There is also enhanced overall drug resistance in hSC31 where even the top-ranked combination of eribulin and OTX015 demonstrated less than 50% killing (Fig 3Bii). To display the interactions that occurred within our panel of chemotherapeutic and targeted agents, a polygonogram was used to illustrate the degree of drug-drug interactions (Fig. 3C). In this analysis, hSC06 was sensitive to a range of pazopanib-based drug combinations, including the combination of OTX015 and pazopanib (Fig. 3Ci). hSC31, in contrast, showed OTX015 and eribulin-based drug combinations to be the most effective, while many of the drug-drug interactions with pazopanib were ablated (Fig. 3Cii). QPOP-derived response surface maps show that pazopanib is synergistic with OTX015 in hSC06 but efficacy of hSC31 is predominantly attributed to efficacy of single agent OTX015 (Fig. 3D). Surprisingly, QPOP analysis also revealed antagonistic interactions within current standard of care drug combinations used against SFT, such as ifosfamide in combination with doxorubicin, where increasing the concentrations of the drugs instead resulted in higher cell viability (Fig. 3Di, 3Dii).

Dose-response drug sensitivity assays were also run separately to deduce the sensitivity of the matched samples towards individual drugs from the QPOP drug panel. Single drug testing of the 12-drug panel revealed pazopanib resistance in hSC31, with IC_50_ 13-fold higher as compared to hSC06 (Fig. S4, Table S2), corroborating clinicoradiological responses in the patient where pazopanib induced response in the liver metastasis (hSC06) but not the brain lesion (hSC31). Interestingly, single drug dose responses of other anti-angiogenic multityrosine kinase inhibitors including sorafenib, sunitinib, lenvatinib and regorafenib also showed a lower IC_50_ than mean across all cancer cell lines in Genomics of Drug Sensitivity (GDSC) database (Fig. S4, Table S2), suggesting that alternative anti-angiogenics could be effective in the pazopanib-resistant brain lesion.

### BET inhibition and pazopanib demonstrates synergistic activity in the liver metastasis

For the initial QPOP analyses, we used data from an OACD with a minimum number of combinations to accurately capture drug dosing. The R^2^ of these QPOP analyses were 0.980 and 0.972 for hSC06 and hSC31 respectively (Fig. S5), indicating the close proximity of the data points to the linear regression analysis. To ascertain quadratic surface mapping accuracy, the QPOP-derived combinations were experimentally verified by dose-response drug sensitivity assays. We determined the IC_50_ of these drugs, either as monotherapies or drug combinations (Fig. 4A). When OTX015 and pazopanib (OTX+PAZO) were administered concurrently at a starting concentration of 100μm in hSC06, there was a marked shift in the dose-response curve, and the IC_50_ of the drug combination decreased compared to when drugs were administered alone (Fig. 4Ai). This indicates that the combination of these two drugs contributed to a strong synergistic increase in efficacy, corroborating RSM data. Although the IC_50_ of OTX015 only saw a moderate reduction from 0.767μM to 0.355μM, the IC_50_ of pazopanib was significantly reduced 80-fold from 28.7μM to 0.355μM. This observation suggests that the efficacy of this top-ranked drug combination is likely due to the increased efficacy of pazopanib in hSC06. On the contrary, when the same combination was administered in hSC31, there minimal shift in the dose-response curve compared to when drugs were administered alone (Fig. 4Aii). This is consistent with the F_a_-CI quantitative diagnostic plot which revealed that co-treatment led to an increasingly synergistic trend (CI < 1) more pronounced in hSC06 compared to hSC31, with larger effect size F_a_. In both hSC06 and hSC31, ifosfamide and doxorubicin were administered concurrently at a starting concentration of 100μM and 10¼M respectively. Dose-response analysis of the ifosfamide and doxorubicin (IFOS+DOX) combination compared to single-drug response confirmed the overall antagonistic interactions within this regimen. Though the IC_50_ of ifosfamide was reduced from 85.4μM to 58.3μM, the efficacy of doxorubicin was reduced as its IC_50_ increased from 0.384μM to 0.583μM with combinatorial treatment in hSC06 (Fig. 4Ai). A similar trend was observed for hSC31 (Fig. 4Aii). This antagonistic interaction was further verified by the F_a_-CI quantitative diagnostic plot which showed to an increasingly antagonistic trend (CI > 1) with larger effect size F_a_. These results collectively provide further evidence that the QPOP-derived optimized combination of OTX+PAZO is more effective than the current standard of care drug combination IFOS+DOX.

**Figure 4.**
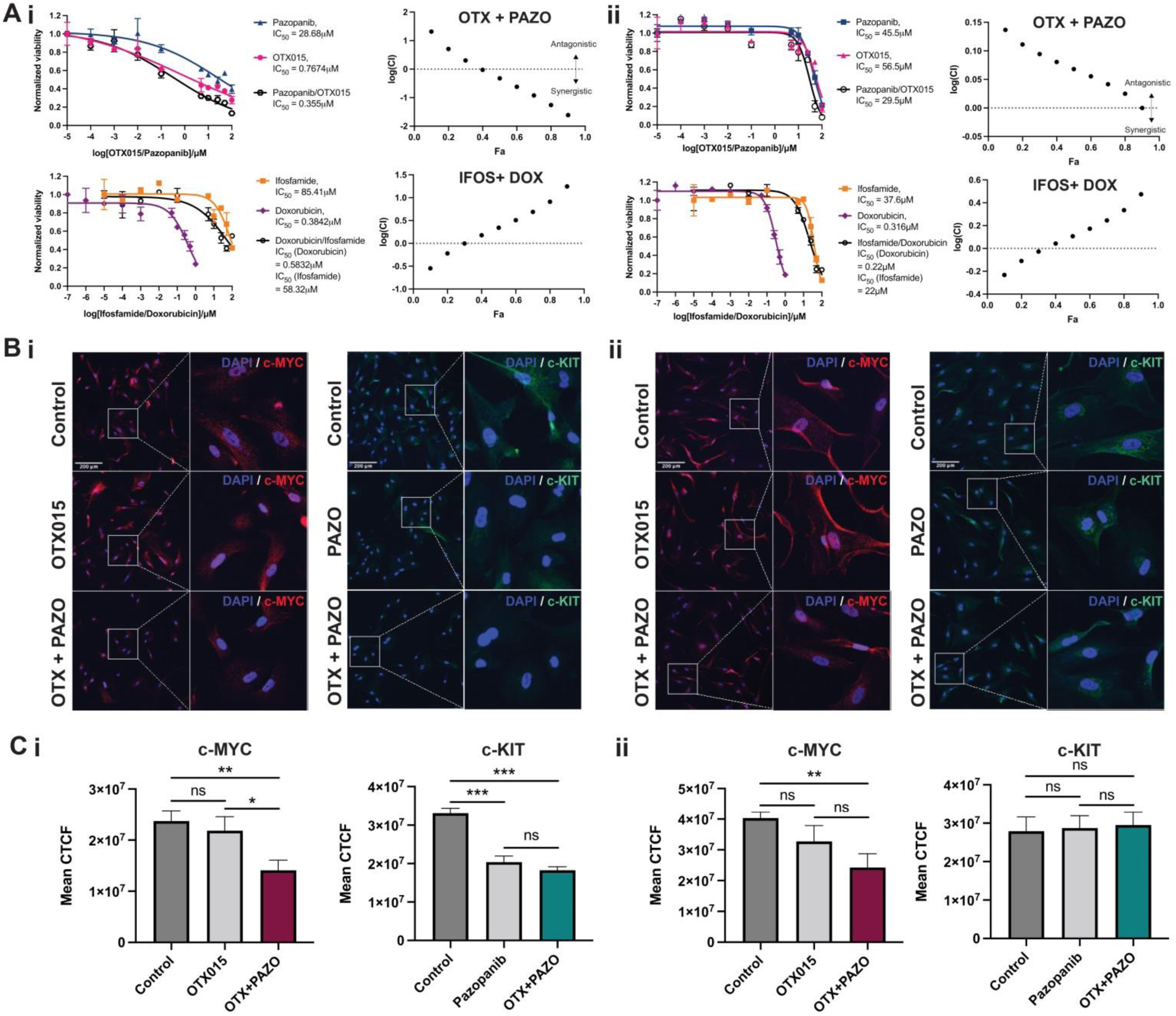
QPOP-derived combination OTX+PAZO demonstrates enhanced synergistic activity compared to standard of care combination IFOS+DOX in the liver metastasis. **(A)** Single drug and combination dose response curves of QPOP-derived drug combination OTX+PAZO and standard of care regimen IFOS+DOX across **(i)** hSC06 and **(ii)** hSC31, with accompanying F_a_-CI (fraction affected-combination index) plot. Combination indices of <1 across a range of effect sizes (F_a_) is indicative of a synergistic interaction. Data are presented as means ± SD of two technical replicates. **(B)** c-MYC (red) or c-KIT (green) is counterstained with DAPI (blue), with accompanying quantification of corrected total cell fluorescence (CTCF) across **(i)** hSC06 and **(ii)** hSC31. Scale bar = 200 μm. Data are means ± SD (n = 3). *P < 0.05; **P < 0.01; ***P < 0.001; ns, not significant (2-tailed Student’s t test).

Further investigation into the effects of this QPOP-derived drug combination revealed tumor specific responses to OTX+PAZO that were not shared across hSC06 and hSC31. BET inhibitor OTX015 has been shown to target BRD2 and BRD4 and decrease c-MYC expression in leukemic cells (38), while the multityrosine kinase activity of pazopanib has been demonstrated in a broad spectrum of solid cancers with potent activity against KIT, PDGR and VEGFR (39). Corroborating dose-response data, c-MYC and c-KIT protein expression were more significantly downregulated in hSC06 than hSC31 following combinatorial treatment, compared to single drug treatments (Fig. 4Bi, 4Ci). This also supports the enhanced efficacy of the OTX+PAZO combination in hSC06 than hSC31, and as predicted by QPOP analysis. Notably, there was no change in c-KIT expression when hSC31 was treated with pazopanib alone (Fig. 4Bii, 4Cii), consistent with pazopanib resistance seen in parallel with single-drug dose-response assay and the patient’s disease progression on pazopanib.

### Transcriptome analysis uncovered a higher expression of apoptosis associated genes following OTX+PAZO treatment in liver metastasis

To explore the molecular basis of treatment response, a comprehensive transcriptomic analysis was performed to evaluate early expression changes. Consistent with their known mechanisms of action, treatment with pazopanib showed a low number of differentially expressed genes, whereas OTX015 showed a high number of differentially expressed genes, compared to controls (Fig. 5A). To understand the molecular effect of combinatorial therapy in hSC06, we performed gene set enrichment analysis [GSEA (40,41), R package ClusterProfiler v4.0.5 (42)] in OTX+PAZO versus control (Fig. 5B). GSEA analysis against the Kyoto Encyclopedia of Genes and Genomes (KEGG) pathways (43) showed downregulation of pathways that may explain cell death with treatment, such as base excision repair, mismatch repair, homologous recombination, DNA replication and cell cycle pathways (Fig S6A). However, these pathways were also downregulated in OTX015 treatment alone (data not shown) and were not specific to combination treatment.

**Figure 5.**
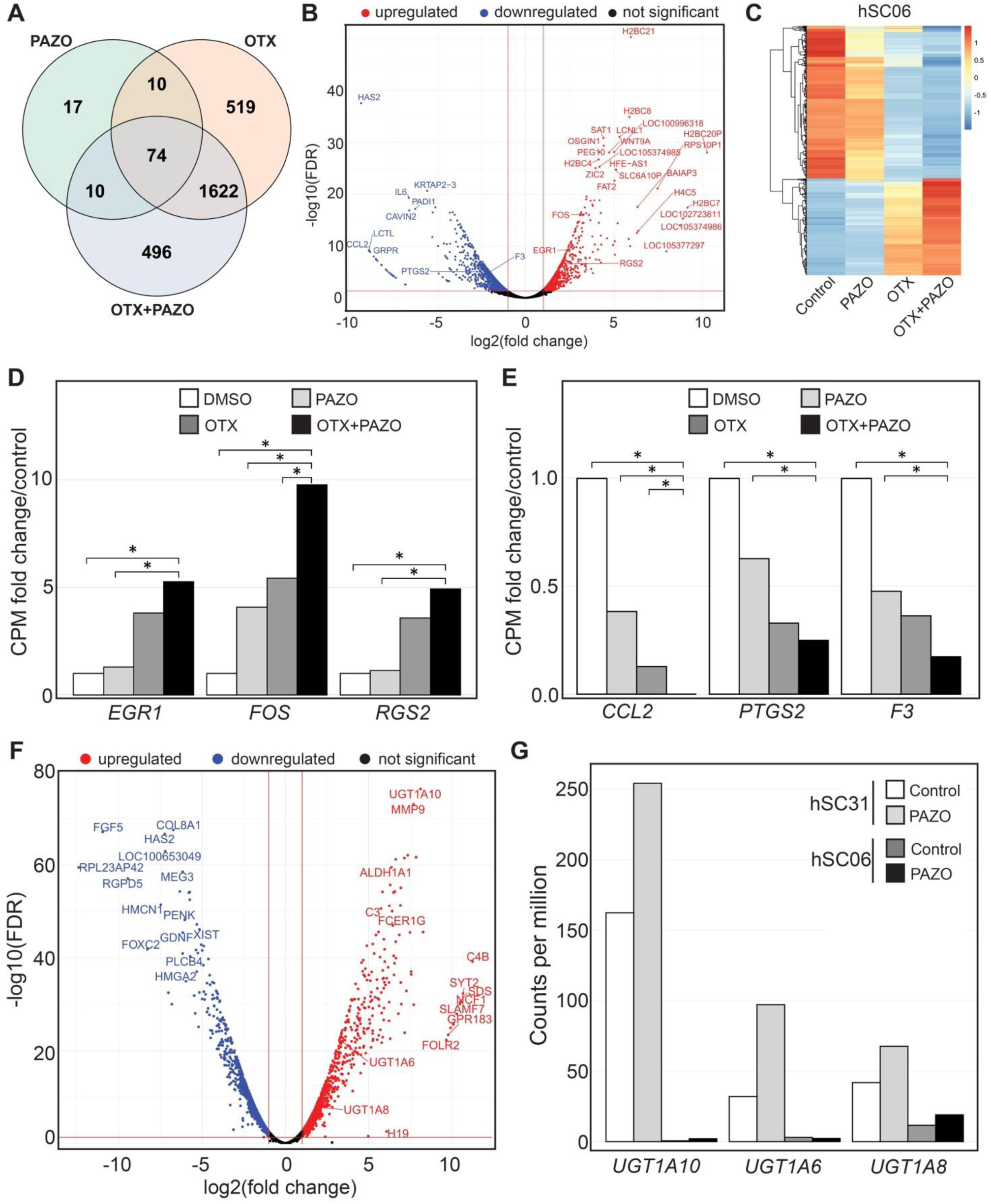
Transcriptome analysis uncovered higher expression of apoptosis associated genes following OTX+PAZO treatment, and highlighted UGT1A10 as a potential biomarker associated with pazopanib resistance. **(A)** A Venn diagram showing the number of overlapping differentially expressed genes (log2foldchange > ±1, false discovery rate (FDR) < 0.05) between PAZO, OTX and OTX+PAZO in hSC06 relative to control. Control is the averaged expression of the untreated and DMSO-treated samples. **(B)** Volcano plot of OTX+PAZO versus control in hSC06. Red vertical lines mark log2foldchange > ±1 and the red horizontal line marks FDR < 0.05. Upregulated genes are coloured red and downregulated genes are coloured blue. **(C)** Heatmap of genes with enhanced up and downregulation in OTX+PAZO treatment relative to the single agents in hSC06. Genes were first filtered for expression changes relative to control in the same direction in both single agents, followed by significant differential expression in OTX+PAZO relative to either single agent in the same direction (p-value < 0.05, log2foldchange> ±1). See Table S4 for the list of genes. Expression values in counts per million (CPM) are scaled row wise. **(D & E)** Expression fold change relative to control of pazopanib downstream effector genes showing enhanced upregulation (FOS, EGR1 and RGS2) and downregulation (CCL2, PTGS2 and F3) in OTX+PAZO relative to the single agents. Fold change in CPM. Asterisk (*) represents p-value < 0.05. **(F)** Volcano plot of differentially expressed genes between hSC31 and hSC06. Red vertical lines mark log2foldchange > ±1 and the red horizontal line marks FDR < 0.05. Upregulated genes are coloured red and downregulated genes are coloured blue. **(G)** Gene expression for three members of the UGT1A UDP glucuronosyltransferase family. Expression is higher in hSC31 compared to hSC06. Gene expression in CPM.

Given that the IC_50_ of pazopanib decreased more than 80-fold when administered in combination with OTX015, we next evaluated the kinase receptor binding targets of pazopanib and their downstream effector genes to see if combination with OTX015 enhanced their change in expression. Pazopanib binding targets showed a mixed response overall with pattern consistency within gene families, although the expression of *KIT* showed more decrease with OTX+PAZO compared to pazopanib alone in hSC06 (Fig. S6B). Next, we postulated that genes involved in the synergy of OTX+PAZO would display enhanced upregulation or downregulation in the same direction, relative to single agent treatment. We first filtered for genes with expression changes in the same direction in both single agent treatments relative to control, followed by genes in OTX+PAZO with significant differential expression in the same direction (p-value < 0.05, log_2_foldchange > ±1) relative to either single agent. This identified a set of 561 candidate genes (Fig. 5C, Table S4). We looked at downstream genes of the kinase receptors of pazopanib in this set of 561 genes and identified a set of early response genes (*FOS, EGR1* and *RGS2*) that showed increased expression upon single-agent treatment and further upregulated with combination treatment (Fig. 5D). Similarly, we also identified three early response genes (*CCL2, PTGS2* and *F3*) that showed decreased expression with single-agent treatment and further downregulated with OTX+PAZO treatment (Fig. 5E).The altered expression of all these genes have been associated with an increased susceptibility to apoptosis (44–52), suggesting that the synergistic cell death observed with OTX+PAZO could be mediated by apoptosis.

### UGT1A glucuronidase family as potential biomarkers associated with pazopanib resistance in SFT

Next, we investigated the molecular basis for clinical progression of the brain recurrence through pazopanib treatment by identifying genes that contribute to pazopanib resistance specific to hSC31. We performed a differential gene analysis between hSC31 and hSC06 (Fig. 5F) and observed that the gene *UGT1A10* showed more than 100-fold higher expression, while *UGT1A6* and *UGT1A8* showed more than 10-fold higher expression in hSC031 (FPKM=84.4), compared to hSC06 (FPKM=0.3)(Fig. 5G). The UGT1A glucuronidase family has been implicated in intrinsic (53,54) and acquired resistance against a range of more than 40 FDA-approved drugs including tyrosine kinase inhibitors like sunitinib and pazopanib (55). Interestingly, treatment of hSC31 with pazopanib increased the expression of *UGT1A10* by 40%. A similar pattern of expression was also seen in other *UGT1A* members, namely *UGT1A6* and *UGT1A8* (Fig. 5G). Taken together, it is possible that the UGT1A family contributes to mediating intrinsic resistance to pazopanib in SFT with corresponding upregulation of expression upon treatment.

### Eribulin sensitivity identified on QPOP provided effective disease control

In keeping with pazopanib resistance seen in hSC31, the patient’s residual brain disease following her last craniotomy had progressed despite pazopanib (Fig. 1A, S2P, S2Q). She had also developed pazopanib-related hypothyroidism and this was corrected with levothyroxine administration. She was then managed expectantly as she was asymptomatic of her disease. Her brain lesion then grew from 1.4cm to 2cm in two months (Fig. 1A, S2Q, S2R) while her extracranial disease remained stable (not shown). Collective decision was made to resume palliative systemic treatment. Because we saw a shift toward eribulin sensitivity in hSC31 and since hSC06 also showed eribulin sensitivity (Fig. 3A, B), the patient was therefore given eribulin for isolated progression of her brain disease, which may also control her disease elsewhere (Fig. 1A, S2). Intriguingly, this halted disease progression in the brain with stable disease extracranially (Fig. 1A, S2Q-S), providing further support that our PDSC can model patient response and QPOP can predict drug response in this setting.

## DISCUSSION

Given the limited treatment options available for unresectable SFT, the identification of effective systemic treatment is an area of unmet need. We showed that PDSCs from this patient with an ultra-rare sarcoma recapitulated the phenotypic expression and behaviour of their matched tumors. Using a QPOP-directed protocol, we systematically screened a 12-drug search set, consisting of 531,441 total possible QPOP-derived combinations and single-agent effects, to identify tumor-specific drug responses in a patient-matched locally recurrent brain lesion and liver metastasis of a rare SFT. This systematic approach identified drug combinations that would outperform clinically-approved regimens, as well as single drug options. Eribulin sensitivity identified from QPOP of her PDSC led to effective disease control.

Doxorubicin is widely applied as first-line therapy for patients with unresectable advanced soft tissue sarcomas including SFTs (56). We noticed that different drug analytic approaches yielded disparate conclusions for doxorubicin. For example, the IC_50_ of doxorubicin in our PDSCs were shown to be comparable to the mean across all doxorubicin-sensitive cell lines on the GDSC database, suggesting sensitivity to doxorubicin. However, liposomal doxorubicin failed to demonstrate disease control in our patient, and this resistance was recapitulated by QPOP in which doxorubicin was ranked low as single agent. This suggests more accurate correlation of QPOP drug ranking than using IC_50_ values from the GDSC database.

Given the angiogenic properties of SFTs, pazopanib has been used for recurrent or metastatic SFT in first- and second-line setting (57). Thus, the patient was given pazopanib as second-line treatment, but showed mixed response in her liver lesions and progression in her dominant brain lesion. This clinical pattern of pazopanib response corroborated with both IC_50_ in PDSC and QPOP analysis. Pazopanib showed an almost 10-fold higher IC_50_ in hSC31 compared to hSC06. Pazopanib as a single agent was ranked low by QPOP assay in hSC31. Notably, progression of the brain lesion on pazopanib treatment cannot be attributed to inability to penetrate the blood brain barrier (BBB). Pazopanib is known to enter an intact BBB (58) and furthermore, the BBB in our patient has been disrupted following multiple craniotomies. Moreover, the IC_50_ of pazopanib in hSC31 was higher compared to mean across all cell lines on GDSC database suggesting that the locally recurrent brain lesion was highly resistant to pazopanib. Interestingly, this may not be true for all anti-angiogenic multityrosine kinase inhibitors, as others such as sorafenib, sunitinib, lenvatinib and regorafenib showed a lower or comparable IC_50_ than the mean across all cancer cell lines in GDSC database. Given that angiogenesis is an important feature involved in tumor growth and metastatic diffusion of SFT, the inhibition of this pathway via other antiangiogenic agents can be considered in our patient.

Intriguingly, single-drug dose-response and QPOP assays highlighted potential tumor response towards eribulin as single agent or in combination, with both brain lesion and liver metastasis displaying shared sensitivity. This finding led to our patient receiving eribulin when she experienced isolated brain progression. As predicted experimentally, eribulin did indeed halt further progression in the brain. Eribulin is a microtubule inhibitor that has been reported to show antitumor activity against soft tissue sarcomas (59). There is retrospective evidence indicating that eribulin exhibits meaningful efficacy as a first-line treatment for soft tissue sarcoma patients in whom doxorubicin is contraindicated (60) and in patient cohorts with liposarcomas in phase III studies (61,62). There is a dearth of evidence for its efficacy in SFT. Notably, eribulin has shown antitumor activity in preclinical xenograft models of SFT (63). Our data may therefore lend support to ongoing clinical trials in SFT cohorts with eribulin.

Additionally, QPOP identified the potential combination of OTX+PAZO with synergistic interactions in hSC06 and prioritized this drug pair over conventional IFOS+DOX which showed no synergy. While the efficacy of pazopanib as a single agent has been largely explored in SFTs, the efficacy of pazopanib in combination with a BET inhibitor has never been investigated in the clinical setting. We postulate that the effect of combinatorial synergy is due to OTX015 enhancing the potency of an already effective pazopanib response as observed from a significant reduction in the IC_50_ of pazopanib when administered with OTX015. Pazopanib is both anti-angiogenic and inhibits multiple tyrosine kinase oncogenic pathways. Previous studies have suggested that synergistically targeting tyrosine kinase signalling and BRD4 may be useful in the treatment of sarcomas. Targeting the IGF1R/PI3K/AKT pathway has been shown to sensitize Ewing Sarcoma to BET bromodomain inhibitors, inducing a strong synergistic response and potent apoptosis in combination both *in vitro* and *in vivo* (64,65). Furthermore, JQ1 has been reported to suppress tumor angiogenesis in sarcoma xenograft models of childhood cancer (66). These results provide proof-of-concept evidence for combining BET inhibitors with pazopanib in this setting.

Although QPOP does not rely on mechanistic assumptions to derive optimal drug combinations, we were able to identify downstream pathways through which OTX+PAZO may act. Common among the three upregulated genes with combination treatment (*FOS, EGR1* and *RGS2*) is previously published data showing their involvement in mediating both proliferative and apoptotic phenotypes (44–47,67,68). We postulate that there might be a context or dosage dependent effect of these genes where in some contexts increase in expression results in proliferation but further overexpression would lead to apoptosis. However this dosage dependent phenotype would need to be validated. Surprisingly, *EGR1* was found to be overexpressed under combinatorial treatment though its overexpression is recognised to be driven by the pathognomonic *NAB2-STAT6* gene fusion. The gene fusion hijacks NAB2 interaction with EGR1 by cleaving off its inhibitory domain and fusing on a transactivating domain from STAT6, altering its function from inhibitory to constitutively active (69). We postulate that overexpression of *EGR1* as a result of combinatorial treatment contributes to increased apoptosis, as shown in other publications which demonstrated the role of EGR1 overexpression in mediating apoptotic cell fates in both neurons and cancers (45,70). The IC_50_ of pazopanib was 10-fold higher in hSC31 as compared to hSC06, we postulate that this is due to the high expression of glucuronidase UGT1A in hSC31 that may be enzymatically modifying pazopanib, leading to reduced binding and inhibition of tyrosine kinase receptors. Future experiments can be performed to test whether selective inhibition of UGT1A might help resensitize the cells to pazopanib.

This study has several limitations. The QPOP assay was performed based on a selected panel of 12 drugs and does not represent all clinically relevant drug candidates, including temozolomide, bevacizumab or other anti-angiogenic agents. Moreover, since drug combinations are administered simultaneously, QPOP does not account for the effect of sequential drug treatment in the combination. In future, it would be crucial to develop pharmacogenomic models which integrate targetable hits derived from “multiomic” data with QPOP-predicted drug combinations to further refine therapeutic predictions for individual patients. We were also unable to functionally validate *UGT1A* candidate genes for their association with pazopanib resistance, given the limited availability of non-immortalised PDSC.

This case is unique as it is the only reported cerebroventricular SFT case in the literature that widely metastasized (Table S3). However, the clinical evolution of any SFT is unpredictable, as local recurrence may occur in 20% to 85% of cases and metastasis in 12% to 36% (71,72). To date, no clinically actionable targets have been identified in SFTs (73). In relapsed or refractory disease, local therapy is given with palliative intent (74), for lack of effective systemic options. QPOP is potentially a solution to systematically identify effective drug combinations in an unbiased manner. By applying experimental data to a deterministic model independent of molecular mechanistic assumptions, QPOP was able to identify clinically actionable regimens against this ultra-rare tumor. Indeed, the identification of eribulin did demonstrate clinical utility. Based on our present findings, options for further treatment can also include other multityrosine kinase inhibitors such as sorafenib, sunitinib, lenvatinib and regorafenib. In future, QPOP-guided treatment can be evaluated in a prospective clinical trial, compared to physician choice of systemic treatment. This truly personalised approach may be highly valuable in the context of rare tumors, such as SFT. Future work is also necessary to uncover the molecular mechanisms that underlie effective drug combinations and to evaluate potential biomarkers for the prediction of tumor responses to specific drugs, for the improvement of patient outcomes in rare cancers.

## CONCLUSIONS

We evaluated therapeutic vulnerability and resistance in a cerebroventricular SFT with recurrence and development of distant metastasis. Concordance between *ex vivo* drug testing results and clinicoradiological response in the patient demonstrates the feasibility of a patient-based drug analytic approach to identify therapeutic options in an ultra-rare soft tissue sarcoma. A novel BETi-pazopanib combination was more effective with enhanced synergy compared to standard of care IFOS+DOX in the metastatic tumor. In SFT, members of the *UGT1A* family are potential biomarkers associated with pazopanib resistance. Eribulin sensitivity identified on QPOP of her PDSC provided effective disease control, suggesting that our PDSC can model patient response and QPOP can predict drug response in this setting. Further work will contribute to the broadening landscape of patient-based drug analytics and catalyze their clinical application in patients with ultra-rare cancers.

## Data Availability

All data produced in the present study are available upon reasonable request to the authors

## List of Abbreviations

*SFT:*: Solitary fibrous tumour
*STS:*: Soft tissue sarcoma
*PDSC:*: Patient-derived SFT cells
*WHO:*: World Health Organization
*QPOP:*: Quadratic Phenotypic Optimization Drug Screening Platform
*OACD:*: Orthogonal array composite design
*IC*_*50*_: half maximal inhibitory concentration
*RSM:*: Response surface mapping
*MRI:*: Magnetic resonance imaging
*CT:*: Computed tomography
*CTCF:*: Corrected total cell fluorescence
*GDSC:*: Genomics of Drug Sensitivity
*FPKM:*: Fragments per kilobase of transcript per million mapped reads
*GSEA*: Gene set enrichment analysis
*F*_*a*_: Fraction affected
*CI:*: Combination Index
*FDR:*: False discovery rate
*CPM:*: Counts per million
*BBB:*: Blood brain barrier

## DECLARATIONS

### Ethics approval and consent to participate

The present study was carried out to comply with the ethical standards of the Helsinki Declaration and approved by the SingHealth Centralized Institution Review Board (CIRB-2018/3182) and the National University of Singapore Institutional Review Board (NUS-IRB-2021-251).

### Consent for publication

Consent for publication has been obtained from the patient.

### Availability of data and materials

Datasets generated and analyzed in this study are included in this published article and its supplementary information files. Raw data and materials are available from the corresponding authors on reasonable request.

### Competing interests

EKC is a shareholder of KYAN Therapeutics.

### Funding

This work was supported by the National Medical Research Council Transition Award (TA20nov-0020), SingHealth Duke-NUS Oncology Academic Clinical Programme (08/FY2020/EX/67-A143 and 08/FY2021/EX/17-A47), the Khoo Pilot Collaborative Award (Duke-NUS-KP(Coll)/2022/0020A), the National Medical Research Council Clinician Scientist-Individual Research Grant-New Investigator Grant (CNIGnov-0025), the National Research Foundation Cancer Science Institute of Singapore RCE Main Grant and the Ministry of Education Academic Research Fund (MOE AcRF Tier 2 [MOE2019-T2-1-115]). CJO is supported by the National Medical Research Council Clinician Scientist-Individual Research Grant (CIRG21jun-0038). VSY is supported by the National Medical Research Council Transition Award (TA20nov-0020).

### Authors’ contributions

VSY, TBT, EKC conceptualized the project. SPYC, BL, BJC, SXX performed experiments and data collection. SPYC, BL, BJC, NS, XYW, TBT, EKC, VSY analyzed the data. VSY, BFL, CJO, CSC, MY, KK, LLO, TKYT contributed to patient management. SPYC, BL, BJC, AV, XYW, TBT, VSY wrote the manuscript. All authors read and approved the final manuscript.

## Acknowledgements

We thank Tan Gek San and Jeremy Ng from the Singapore General Hospital Translational Pathology Centre and the National Cancer Centre Singapore IMPACT team for Oncomine information. We are indebted to our patient and her family for their generous support and contribution towards this work.

## Notes

### Clinical Trial

NCT04986748

### Author Declarations

Ethics committee of the SingHealth Centralized Institution Review Board (CIRB-2018/3182) and the National University of Singapore Institutional Review Board (NUS-IRB-2021-251) gave ethical approval for this work

