## Supplementary Data for "*Ex vivo* drug testing in an ultra-rare sarcoma reveals therapeutic vulnerability and resistance"

**SUPPLEMENTARY INFORMATION**

### Figure S1

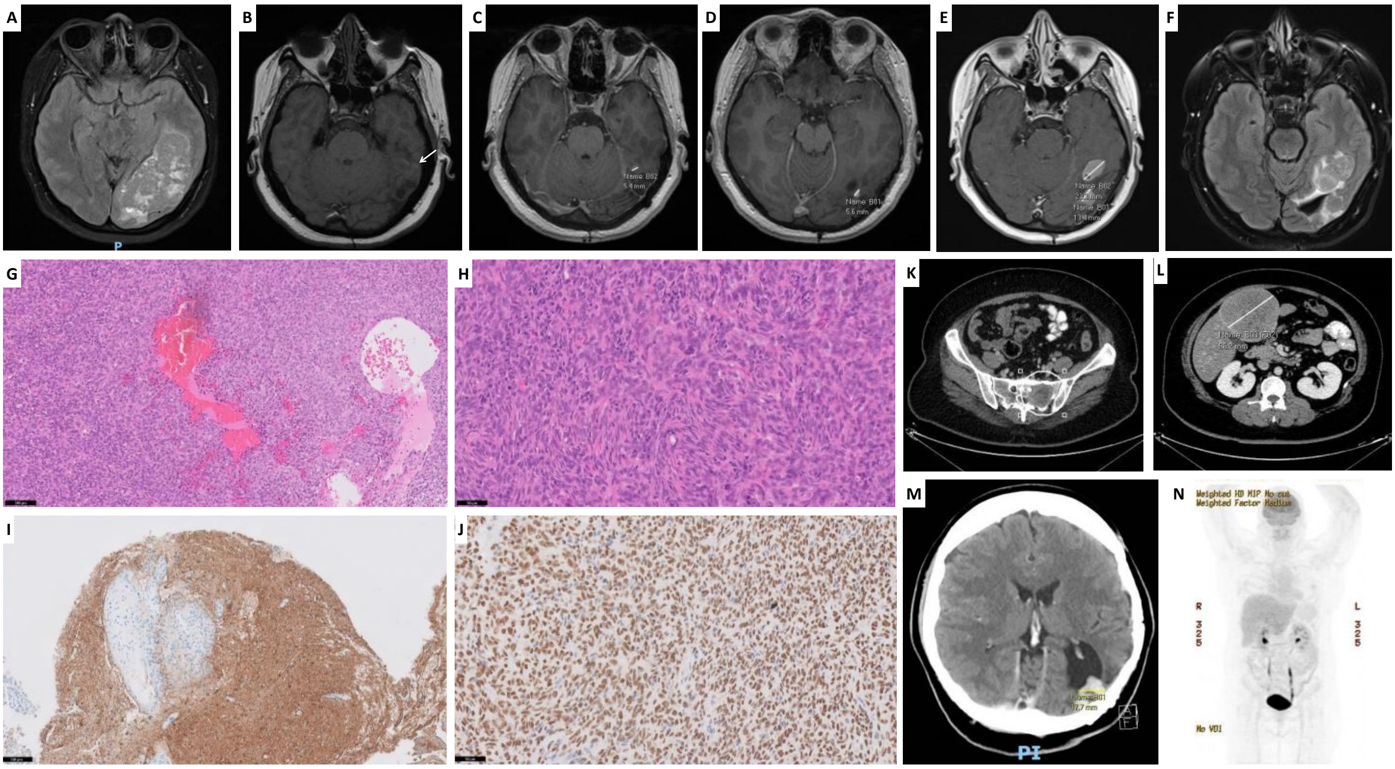


**Figure S1: Clinical course from diagnosis to local recurrence and development of distant metastasis.** Sequential Magnetic resonance imaging (MRI) brain **(A-F)**, photomicrographs of specimen from second craniotomy **(G-J)** and computed tomography (CT) imaging **(K-M)** and positron emission tomography (PET) CT **(N)** at development of distant metastasis are shown. **(A)** Left parieto-occipital mass with a perilesional cerebrospinal fluid cleft suggesting an intraventricular location. An associated 9 mm shift of midline structures with prominence of the right lateral ventricle suggests early hydrocephalus. **(B)** Increase in size of an enhancing focus along the lateral margin of the left occipital horn, marked with an arrow, suggesting local recurrence. **(C, D)** New 0.5 cm nodules were deemed indeterminate. **(E)** Tumour recurrence with enhancing nodules lateral to the surgical cavity at the left occipital horn. **(F)** Further increase in size of recurrent tumour. This was excised. **(G-H)** Haematoxylin and eosin stained section of the brain mass demonstrating markedly cellular sheets of spindled tumour cells arranged in ill-defined fascicles, with only small amounts of collagen, as well as “staghorn” vessels (G – 100x magnification, 10x objective, H – 200x magnification, 20x objective). **(I)** GFAP immunoperoxidase stained section of the brain mass highlighting cortical invasion (100x magnification, 10x objective). **(J)** STAT6 immunoperoxidase stained section of the brain mass showing diffuse nuclear immunoreactivity (200x magnification, 20x objective). **(K)** Bone metastases in the left hemisacrum and iliac bone. **(L)** Dominant liver metastasis measuring 9.6 cm. **(M)** Largest left parieto-occipital lesion measuring 1.7 cm. **(N)** Fluorodeoxyglucose (FDG) avidity of all lesions was low.

### Figure S2

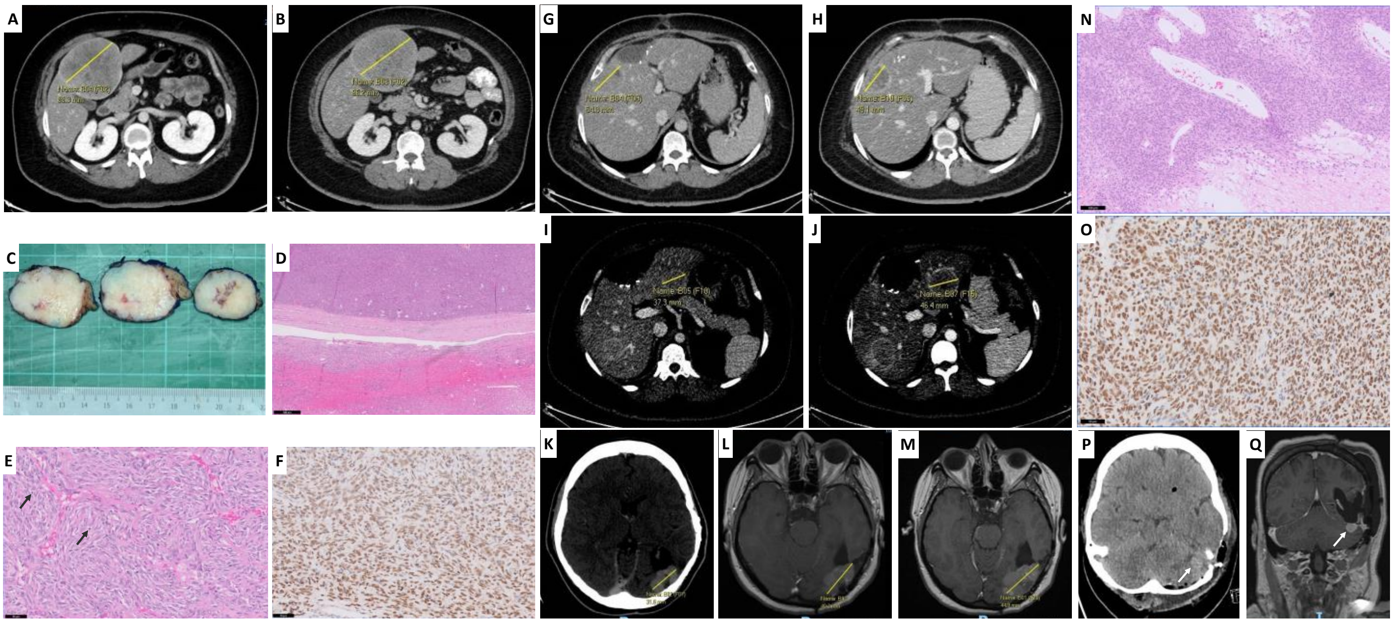


**Figure S2: Clinical course following initiation of systemic treatment**. **(A-B)** CT imaging showing progression of the dominant liver metastasis on Liposomal Doxorubicin from 8.9 cm to 9.6 cm. This lesion was resected. **(C)** Gross specimen photograph of the liver resection specimen, showing a tumour present within the liver parenchyma. **(D-E)** Haematoxylin and eosin stained sections of the liver mass demonstrating a tumour comprised of cellular sheets of spindled tumour cells arranged in ill-defined fascicles, with only small amounts of collagen, as well as “staghorn” vessels arising from the liver, with increased mitotic figures, marked with arrows (D – 20x magnification, 2x objective, E – 200x magnification, 20x objective). **(F)** STAT6 immunoperoxidase stained section of the liver mass showing diffuse nuclear immunoreactivity (200x magnification, 20x objective). **(G-H)** CT imaging showing response in the marked liver metastasis with Pazopanib treatment, decreasing in size from 5.5 cm to 4.9cm. **(I-J)** In a separate liver metastasis, there was increase in size from 3.7 cm to 4.6 cm with Pazopanib treatment. **(K-M)** Brain imaging showing continued increase in size of the dominant brain lesion through Pazopanib treatment. This was resected during her third craniotomy. **(N)** Haematoxylin and eosin stained section of the brain mass from her third craniotomy demonstrating markedly cellular sheets of spindled tumour cells arranged in ill-defined fascicles, with only small amounts of collagen, as well as dilated vessels and areas of necrosis (100x magnification, 10x objective). **(O)** STAT6 immunoperoxidase stained section of the brain mass showing diffuse nuclear immunoreactivity (100x magnification, 10x objective). **(P)** CT brain one day post-operatively showing residual tumour, marked with an arrow. This lesion was not seen intraoperatively. **(Q)** MRI Brain two months later showing a 1.4 cm residual lesion, marked with an arrow.

### Figure S3

**A**

Assay & Sample Information:

Date of Consent: [redacted]

Sample Type: FFPE, Left parieto-occipital tumour

Tumour Percentage/Area: >95% / 546$mm^2$

Procedure Date: [redacted]

Sequencing Performance Specification:

| *DNA sequencing* | *RNA sequencing* |
|---|---|
| On-target reads: 34880578 (95.87%) | On-target reads: 249227 (62.81%) |
| Average base coverage depth: 2806.24 | Average base coverage depth: 80.11 |
| Coverage: 97.53%(250X), 95.15%(500X) | Max. coverage depth: 48865 |

| |
|---|
| **Amplification[1] (number of copies)**<br>NIL |
| **Fusion**<br>* Poor RNA quality. No coverage. |
| **Tumour Mutation Burden (TMB)**<br>* The estimated mutation burden = 2.84 mutations per Mbp |
| **Microsatellite Instability (MSI)**<br>MSI status/score: MSS / 3.71 (MSS < or = 20; MSI-H > 20) |
| **Other variants[2]**<br>ARHGAP35, NM_004491, Exon1, c.266G>A, p.Cys89Tyr (p.C89Y)<br>DPYD, NM_000110, Exon12, c.1519G>A, p.Val507Ile (p.V507I)<br>ERBB3, NM_001982, Exon13, c.1493A>T, p.Lys498Ile (p.K498I)<br>ERCC2, NM_000400, Exon10, c.934G>A, p.Asp312Asn (p.D312N)<br>FANCI, NM_018193, Exon28, c.2999T>C, p.Ile1000Thr (p.I1000T)<br>KMT2D, NM_003482, Exon31, c.6409A>C, p.Thr2137Pro (p.T2137P)<br>PARP1, NM_001618, Exon14, c.2071-3C>T, (p.?)<br>POLD1, NM_002691, Exon5, c.506A>G, p.Asn169Ser (p.N169S)<br>PRDM1, NM_001198, Exon2, c.239A>T, p.Glu80Val (p.E80V)<br>RNF43, NM_017763, Exon9, c.1165C>A, p.Arg389Ser (p.R389S)<br>SETD2, NM_014159, Exon3, c.590C>T, p.Ala197Val (p.A197V)<br>ZNF217, NM_006526, Exon3, c.2666A>G, p.Asp889Gly (p.D889G)<br>ZNF479, NM_033273, Exon5, c.1129G>A, p.Gly377Arg (p.G377R) |
| **Copy Number Loss (number of copies)**<br>NOTCH1 (1.56), HLA-A (1.25), HLA-B (1.22) |

[1]Amplification calls are made with 4 or more copies of the gene

[2]This is a list of uncurated and unannotated variants detected. For further discussion at IMPACT tumour board

**B**

Assay & Sample Information:

Date of Consent: [redacted]

Sample Type: FFPE, Liver segment 4B resection

Tumour Percentage/Area: 90% / 229$mm^2$

Procedure Date: [redacted]

Sequencing Performance Specification:

| *DNA sequencing* | *RNA sequencing* |
|---|---|
| On-target reads: 26900069 (88.68%) | On-target reads: 666127(85.09%) |
| Average base coverage depth: 2176.24 | Average base coverage depth: 440.62 |
| Coverage: 97.79%(250X), 94.29%(500X) | Max. coverage depth: 84775 |

| |
|---|
| **Amplification[1] (number of copies)**<br>NIL |
| **Fusion**<br>Suboptimal RNA quality. No detectable fusion.<br>Comment: NAB2-STAT6 fusion seen in solitary fibrous tumours is not covered by this assay. |
| **Tumour Mutation Burden (TMB)[2]**<br>* The estimated mutation burden = 5.67 mutations per Mbp |
| **Other variants[3]**<br>ARHGAP35 (NM_004491) Exon1, c.266G>A/p.Cys89Tyr (p.C89Y)<br>DPYD (NM_000110) Exon12, c.1519G>A/p.Val507Ile (p.V507I)<br>ERBB3 (NM_001982) Exon13, c.1493A>T/p.Lys498Ile (p.K498I)<br>ERCC2 (NM_000400) Exon10, c.934G>A/p.Asp312Asn (p.D312N)<br>ERRFI1 (NM_018948) Exon4, c.325G>A/p.Asp109Asn (p.D109N)<br>FANCI (NM_018193) Exon28, c.2999T>C/p.Ile1000Thr (p.I1000T)<br>KDR (NM_002253) Exon11, c.1444T>C/p.Cys482Arg (p.C482R)<br>KMT2D (NM_003482) Exon31, c.6409A>C/p.Thr2137Pro (p.T2137P)<br>PARP1 (NM_001618) Exon14, c.2071-3C>T/. (p.?)<br>POLD1 (NM_002691) Exon5, c.506A>G/p.Asn169Ser (p.N169S)<br>PRDM1 (NM_001198) Exon2, c.239A>T/p.Glu80Val (p.E80V)<br>SETD2 (NM_014159) Exon3, c.590C>T/p.Ala197Val (p.A197V) |
| **Copy Number Loss (number of copies)**<br>CSMD3 (1.54) |

[1]Amplification calls are made with 4 or more copies of the gene

[2]The mutation burden is calculated based on the size of the panel and number of non-synonymous mutations and mathematically approximated for 1Mbp.

[3]This is a list of uncurated and unannotated variants detected. For further discussion at IMPACT tumour board

**Figure S3: Oncomine analysis did not reveal clinically actionable findings.** Oncomine Comprehensive Panel v3 (RNA) and Oncomine Comprehensive Assay Plus (DNA) assays performed on formalin-fixed paraffin-embedded sample (FFPE) from **(A)** initial resection of primary SFT in the left parieto-occipital region of the brain and **(B)** resection of metastatic liver segment 4B lesion.

**Figure S4**

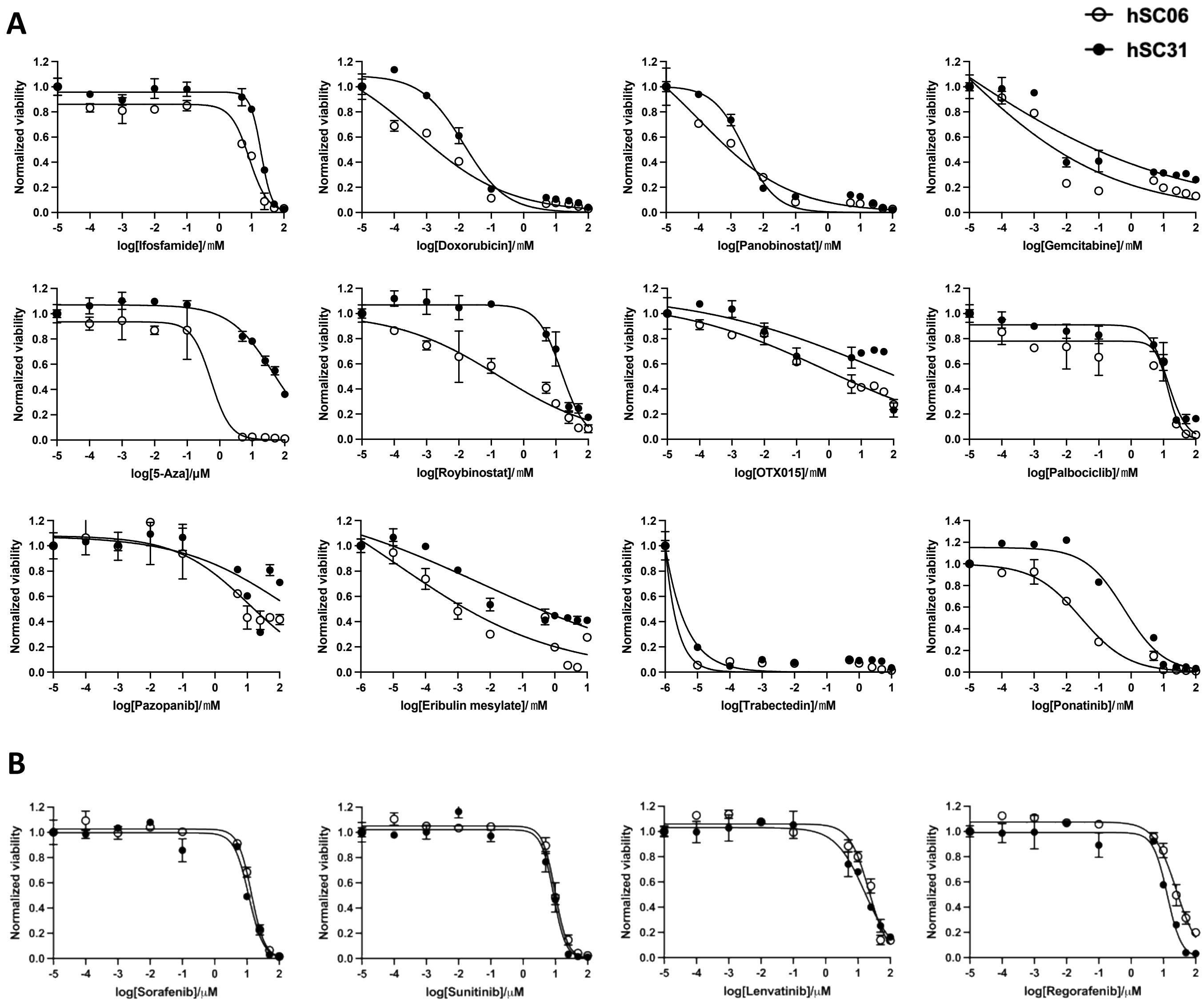


**Figure S4: Single-drug dose-response curves of drug panel.** Single drug dose-response curves of **(A)** all 12 drugs from QPOP screen and **(B)** other clinically used anti-angiogenic agents in hSC06 and hSC31. All dose-response curves are represented as means ± SD of two technical replicates.

### Figure S5

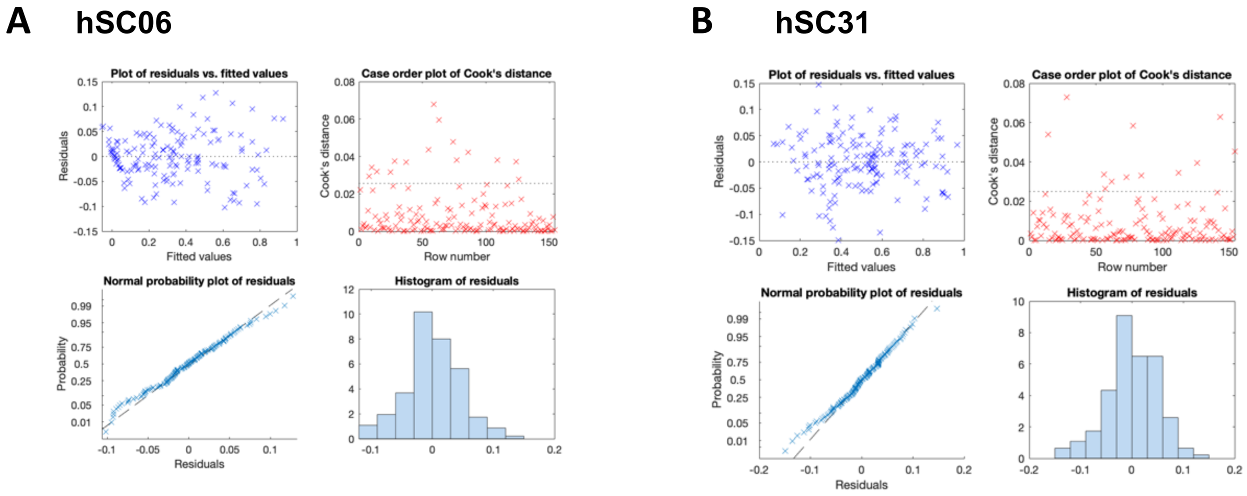


- Experimental data set follows a normal distribution.
- Fitting Correlation = **0.98**
- Z’ Factor = **0.89**
- Strictly Standardized Mean Difference (SSMD; β) = **28.2**

- Experimental data set follows a normal distribution.
- Fitting Correlation = **0.972**
- Z’ Factor = **0.735**
- Strictly Standardized Mean Difference (SSMD; β) = **12**

**Figure S5: QPOP linear regression analysis**. To confirm the fidelity of QPOP, correlation coefficients (measures of the strength of the linear association between two variables ranging in value between zero and one) across **(A)** hSC06 and **(B)** hSC31 were calibrated using the data points in dose response assay of single drugs and drug combination. Z’ score was calculated to ensure the robustness and integrity of the assay.

### Figure S6

**A**

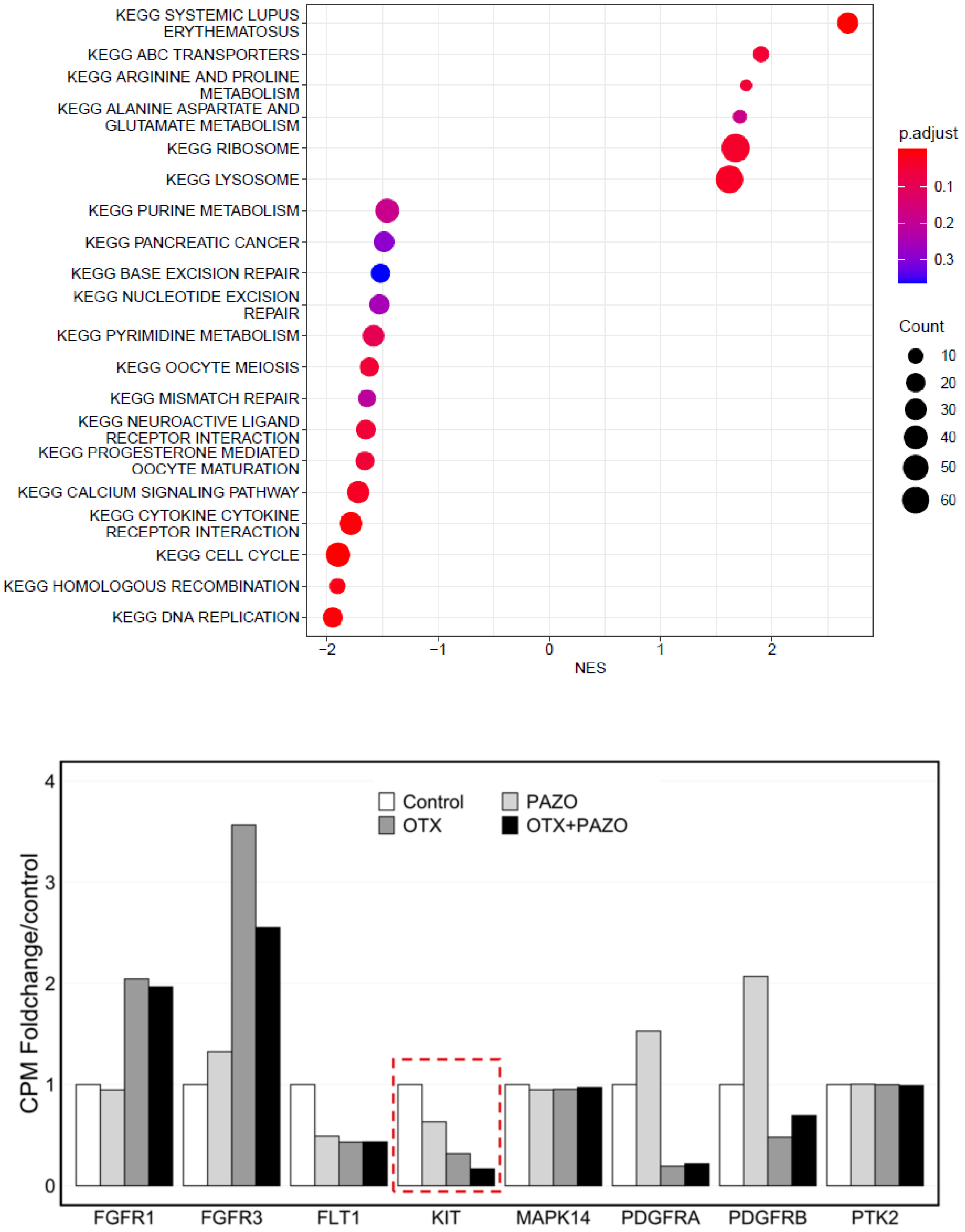


**B**



**Figure S6: Transcriptomic analysis of hSC06 (A)** Dotplot of top 20 Kyoto Encyclopedia of Genes and Genomes (KEGG) pathways enriched through gene set enrichment analysis (GSEA). GSEA was performed for genes differentially expressed in OTX+PAZO relative to control in hSC06. The genes were ranked using their $\log_2$ foldchange for OTX+PAZO versus control. **(B)** Relative expression of pazopanib targeted receptors in hSC06. Most receptors showed mixed expression trends but *KIT* (highlighted in red) was downregulated in OTX, PAZO, and further downregulated in OTX+PAZO. Fold change in counts per million.

**Table S1:** QPOP drug combination design for 12 drugs at three dosage levels (-1, 0, 1) consisting of 155 combinations. The 12 drugs include Ifosfamide (Ifos), Doxorubicin (Doxo), Panobinostat (Pano), Gemcitabine (Gem), 5-Azacytidine (Aza), Roybinostat (Roy), OTX015 (OTX), Palbociclib (Palbo), Pazopanib (Pazo), Eribulin mesylate (Eri), Trabectedin (Tra) and Ponatinib (Pona).

| Run | Ifos | Doxo | Pano | Gem | Aza | Roy | OTX | Palbo | Pazo | Eri | Tra | Pona |
|---|---|---|---|---|---|---|---|---|---|---|---|---|
| 1 | -1 | -1 | -1 | -1 | -1 | -1 | -1 | -1 | -1 | -1 | -1 | -1 |
| 2 | -1 | -1 | -1 | -1 | -1 | -1 | 1 | 1 | 1 | 1 | 1 | 1 |
| 3 | -1 | -1 | -1 | -1 | -1 | 1 | -1 | 1 | -1 | -1 | 1 | -1 |
| 4 | -1 | -1 | -1 | -1 | -1 | 1 | 1 | -1 | 1 | 1 | -1 | 1 |
| 5 | -1 | -1 | -1 | -1 | 1 | -1 | -1 | -1 | -1 | 1 | -1 | 1 |
| 6 | -1 | -1 | -1 | -1 | 1 | -1 | 1 | 1 | 1 | -1 | 1 | -1 |
| 7 | -1 | -1 | -1 | -1 | 1 | 1 | -1 | 1 | 1 | -1 | -1 | 1 |
| 8 | -1 | -1 | -1 | -1 | 1 | 1 | 1 | -1 | -1 | 1 | 1 | -1 |
| 9 | -1 | -1 | -1 | 1 | -1 | -1 | -1 | -1 | 1 | -1 | 1 | 1 |
| 10 | -1 | -1 | -1 | 1 | -1 | -1 | 1 | 1 | -1 | 1 | -1 | -1 |
| 11 | -1 | -1 | -1 | 1 | -1 | 1 | -1 | 1 | 1 | 1 | 1 | -1 |
| 12 | -1 | -1 | -1 | 1 | -1 | 1 | 1 | -1 | -1 | -1 | -1 | 1 |
| 13 | -1 | -1 | -1 | 1 | 1 | -1 | -1 | 1 | -1 | -1 | 1 | 1 |
| 14 | -1 | -1 | -1 | 1 | 1 | -1 | 1 | -1 | 1 | 1 | -1 | -1 |
| 15 | -1 | -1 | -1 | 1 | 1 | 1 | -1 | -1 | 1 | -1 | -1 | -1 |
| 16 | -1 | -1 | -1 | 1 | 1 | 1 | 1 | 1 | -1 | 1 | 1 | 1 |
| 17 | -1 | -1 | 1 | -1 | -1 | -1 | -1 | 1 | -1 | 1 | 1 | 1 |
| 18 | -1 | -1 | 1 | -1 | -1 | -1 | 1 | -1 | 1 | -1 | -1 | -1 |
| 19 | -1 | -1 | 1 | -1 | -1 | 1 | -1 | -1 | 1 | -1 | 1 | 1 |
| 20 | -1 | -1 | 1 | -1 | -1 | 1 | 1 | 1 | -1 | 1 | -1 | -1 |
| 21 | -1 | -1 | 1 | -1 | 1 | -1 | -1 | 1 | 1 | 1 | -1 | -1 |
| 22 | -1 | -1 | 1 | -1 | 1 | -1 | 1 | -1 | -1 | -1 | 1 | 1 |
| 23 | -1 | -1 | 1 | -1 | 1 | 1 | -1 | -1 | 1 | 1 | 1 | -1 |
| 24 | -1 | -1 | 1 | -1 | 1 | 1 | 1 | 1 | -1 | -1 | -1 | 1 |
| 25 | -1 | -1 | 1 | 1 | -1 | -1 | -1 | -1 | -1 | 1 | 1 | -1 |
| 26 | -1 | -1 | 1 | 1 | -1 | -1 | 1 | 1 | 1 | -1 | -1 | 1 |
| 27 | -1 | -1 | 1 | 1 | -1 | 1 | -1 | 1 | 1 | 1 | -1 | 1 |
| 28 | -1 | -1 | 1 | 1 | -1 | 1 | 1 | -1 | -1 | -1 | 1 | -1 |
| 29 | -1 | -1 | 1 | 1 | 1 | -1 | -1 | 1 | -1 | -1 | -1 | -1 |
| 30 | -1 | -1 | 1 | 1 | 1 | -1 | 1 | -1 | 1 | 1 | 1 | 1 |
| 31 | -1 | -1 | 1 | 1 | 1 | 1 | -1 | -1 | -1 | 1 | -1 | 1 |
| 32 | -1 | -1 | 1 | 1 | 1 | 1 | 1 | 1 | 1 | -1 | 1 | -1 |
| 33 | -1 | 1 | -1 | -1 | -1 | -1 | -1 | -1 | 1 | 1 | 1 | -1 |
| 34 | -1 | 1 | -1 | -1 | -1 | -1 | 1 | 1 | -1 | -1 | -1 | 1 |
| 35 | -1 | 1 | -1 | -1 | -1 | 1 | -1 | 1 | -1 | 1 | -1 | 1 |
| 36 | -1 | 1 | -1 | -1 | -1 | 1 | 1 | -1 | 1 | -1 | 1 | -1 |
| 37 | -1 | 1 | -1 | -1 | 1 | -1 | -1 | 1 | -1 | 1 | 1 | -1 |
| 38 | -1 | 1 | -1 | -1 | 1 | -1 | 1 | -1 | 1 | -1 | -1 | 1 |
| 39 | -1 | 1 | -1 | -1 | 1 | 1 | -1 | -1 | -1 | -1 | 1 | 1 |
| 40 | -1 | 1 | -1 | -1 | 1 | 1 | 1 | 1 | 1 | 1 | -1 | -1 |
| 41 | -1 | 1 | -1 | 1 | -1 | -1 | -1 | 1 | 1 | -1 | -1 | -1 |

| | | | | | | | | | | | | |
|---|---|---|---|---|---|---|---|---|---|---|---|---|
| 42 | -1 | 1 | -1 | 1 | -1 | -1 | 1 | -1 | -1 | 1 | 1 | 1 |
| 43 | -1 | 1 | -1 | 1 | -1 | 1 | -1 | -1 | -1 | 1 | -1 | -1 |
| 44 | -1 | 1 | -1 | 1 | -1 | 1 | 1 | 1 | 1 | -1 | 1 | 1 |
| 45 | -1 | 1 | -1 | 1 | 1 | -1 | -1 | 1 | 1 | 1 | -1 | 1 |
| 46 | -1 | 1 | -1 | 1 | 1 | -1 | 1 | -1 | -1 | -1 | 1 | -1 |
| 47 | -1 | 1 | -1 | 1 | 1 | 1 | -1 | -1 | 1 | 1 | 1 | 1 |
| 48 | -1 | 1 | -1 | 1 | 1 | 1 | 1 | 1 | -1 | -1 | -1 | -1 |
| 49 | -1 | 1 | 1 | -1 | -1 | -1 | -1 | -1 | 1 | 1 | -1 | 1 |
| 50 | -1 | 1 | 1 | -1 | -1 | -1 | 1 | 1 | -1 | -1 | 1 | -1 |
| 51 | -1 | 1 | 1 | -1 | -1 | 1 | -1 | 1 | 1 | -1 | -1 | -1 |
| 52 | -1 | 1 | 1 | -1 | -1 | 1 | 1 | -1 | -1 | 1 | 1 | 1 |
| 53 | -1 | 1 | 1 | -1 | 1 | -1 | -1 | 1 | 1 | -1 | 1 | 1 |
| 54 | -1 | 1 | 1 | -1 | 1 | -1 | 1 | -1 | -1 | 1 | -1 | -1 |
| 55 | -1 | 1 | 1 | -1 | 1 | 1 | -1 | -1 | -1 | -1 | -1 | -1 |
| 56 | -1 | 1 | 1 | -1 | 1 | 1 | 1 | 1 | 1 | 1 | 1 | 1 |
| 57 | -1 | 1 | 1 | 1 | -1 | -1 | -1 | -1 | -1 | -1 | -1 | 1 |
| 58 | -1 | 1 | 1 | 1 | -1 | -1 | 1 | 1 | 1 | 1 | 1 | -1 |
| 59 | -1 | 1 | 1 | 1 | -1 | 1 | -1 | 1 | -1 | -1 | 1 | 1 |
| 60 | -1 | 1 | 1 | 1 | -1 | 1 | 1 | -1 | 1 | 1 | -1 | -1 |
| 61 | -1 | 1 | 1 | 1 | 1 | -1 | -1 | -1 | 1 | -1 | 1 | -1 |
| 62 | -1 | 1 | 1 | 1 | 1 | -1 | 1 | 1 | -1 | 1 | -1 | 1 |
| 63 | -1 | 1 | 1 | 1 | 1 | 1 | -1 | 1 | -1 | 1 | 1 | -1 |
| 64 | -1 | 1 | 1 | 1 | 1 | 1 | 1 | -1 | 1 | -1 | -1 | 1 |
| 65 | 1 | -1 | -1 | -1 | -1 | -1 | -1 | 1 | 1 | 1 | -1 | -1 |
| 66 | 1 | -1 | -1 | -1 | -1 | -1 | 1 | -1 | -1 | -1 | 1 | 1 |
| 67 | 1 | -1 | -1 | -1 | -1 | 1 | -1 | -1 | -1 | 1 | 1 | 1 |
| 68 | 1 | -1 | -1 | -1 | -1 | 1 | 1 | 1 | 1 | -1 | -1 | -1 |
| 69 | 1 | -1 | -1 | -1 | 1 | -1 | -1 | -1 | 1 | -1 | 1 | -1 |
| 70 | 1 | -1 | -1 | -1 | 1 | -1 | 1 | 1 | -1 | 1 | -1 | 1 |
| 71 | 1 | -1 | -1 | -1 | 1 | 1 | -1 | 1 | 1 | 1 | 1 | 1 |
| 72 | 1 | -1 | -1 | -1 | 1 | 1 | 1 | -1 | -1 | -1 | -1 | -1 |
| 73 | 1 | -1 | -1 | 1 | -1 | -1 | -1 | -1 | 1 | 1 | -1 | 1 |
| 74 | 1 | -1 | -1 | 1 | -1 | -1 | 1 | 1 | -1 | -1 | 1 | -1 |
| 75 | 1 | -1 | -1 | 1 | -1 | 1 | -1 | 1 | -1 | -1 | -1 | 1 |
| 76 | 1 | -1 | -1 | 1 | -1 | 1 | 1 | -1 | 1 | 1 | 1 | -1 |
| 77 | 1 | -1 | -1 | 1 | 1 | -1 | -1 | -1 | -1 | 1 | 1 | -1 |
| 78 | 1 | -1 | -1 | 1 | 1 | -1 | 1 | 1 | 1 | -1 | -1 | 1 |
| 79 | 1 | -1 | -1 | 1 | 1 | 1 | -1 | 1 | -1 | 1 | -1 | -1 |
| 80 | 1 | -1 | -1 | 1 | 1 | 1 | 1 | -1 | 1 | -1 | 1 | 1 |
| 81 | 1 | -1 | 1 | -1 | -1 | -1 | -1 | 1 | -1 | -1 | -1 | 1 |
| 82 | 1 | -1 | 1 | -1 | -1 | -1 | 1 | -1 | 1 | 1 | 1 | -1 |
| 83 | 1 | -1 | 1 | -1 | -1 | 1 | -1 | -1 | -1 | 1 | -1 | -1 |
| 84 | 1 | -1 | 1 | -1 | -1 | 1 | 1 | 1 | 1 | -1 | 1 | 1 |
| 85 | 1 | -1 | 1 | -1 | 1 | -1 | -1 | -1 | 1 | -1 | -1 | 1 |
| 86 | 1 | -1 | 1 | -1 | 1 | -1 | 1 | 1 | -1 | 1 | 1 | -1 |
| 87 | 1 | -1 | 1 | -1 | 1 | 1 | -1 | 1 | -1 | -1 | 1 | -1 |
| 88 | 1 | -1 | 1 | -1 | 1 | 1 | 1 | -1 | 1 | 1 | -1 | 1 |
| 89 | 1 | -1 | 1 | 1 | -1 | -1 | -1 | 1 | 1 | -1 | 1 | -1 |

| | | | | | | | | | | | | |
|---|---|---|---|---|---|---|---|---|---|---|---|---|
| 90 | 1 | -1 | 1 | 1 | -1 | -1 | 1 | -1 | -1 | 1 | -1 | 1 |
| 91 | 1 | -1 | 1 | 1 | -1 | 1 | -1 | -1 | 1 | -1 | -1 | -1 |
| 92 | 1 | -1 | 1 | 1 | -1 | 1 | 1 | 1 | -1 | 1 | 1 | 1 |
| 93 | 1 | -1 | 1 | 1 | 1 | -1 | -1 | 1 | 1 | 1 | 1 | 1 |
| 94 | 1 | -1 | 1 | 1 | 1 | -1 | 1 | -1 | -1 | -1 | -1 | -1 |
| 95 | 1 | -1 | 1 | 1 | 1 | 1 | -1 | -1 | -1 | -1 | 1 | 1 |
| 96 | 1 | -1 | 1 | 1 | 1 | 1 | 1 | 1 | 1 | 1 | -1 | -1 |
| 97 | 1 | 1 | -1 | -1 | -1 | -1 | -1 | 1 | 1 | -1 | 1 | 1 |
| 98 | 1 | 1 | -1 | -1 | -1 | -1 | 1 | -1 | -1 | 1 | -1 | -1 |
| 99 | 1 | 1 | -1 | -1 | -1 | 1 | -1 | -1 | 1 | -1 | -1 | 1 |
| 100 | 1 | 1 | -1 | -1 | -1 | 1 | 1 | 1 | -1 | 1 | 1 | -1 |
| 101 | 1 | 1 | -1 | -1 | 1 | -1 | -1 | 1 | -1 | -1 | -1 | -1 |
| 102 | 1 | 1 | -1 | -1 | 1 | -1 | 1 | -1 | 1 | 1 | 1 | 1 |
| 103 | 1 | 1 | -1 | -1 | 1 | 1 | -1 | -1 | 1 | 1 | -1 | -1 |
| 104 | 1 | 1 | -1 | -1 | 1 | 1 | 1 | 1 | -1 | -1 | 1 | 1 |
| 105 | 1 | 1 | -1 | 1 | -1 | -1 | -1 | 1 | -1 | 1 | 1 | 1 |
| 106 | 1 | 1 | -1 | 1 | -1 | -1 | 1 | -1 | 1 | -1 | -1 | -1 |
| 107 | 1 | 1 | -1 | 1 | -1 | 1 | -1 | -1 | -1 | -1 | 1 | -1 |
| 108 | 1 | 1 | -1 | 1 | -1 | 1 | 1 | 1 | 1 | 1 | -1 | 1 |
| 109 | 1 | 1 | -1 | 1 | 1 | -1 | -1 | -1 | -1 | -1 | -1 | 1 |
| 110 | 1 | 1 | -1 | 1 | 1 | -1 | 1 | 1 | 1 | 1 | 1 | -1 |
| 111 | 1 | 1 | -1 | 1 | 1 | 1 | -1 | 1 | 1 | -1 | 1 | -1 |
| 112 | 1 | 1 | -1 | 1 | 1 | 1 | 1 | -1 | -1 | 1 | -1 | 1 |
| 113 | 1 | 1 | 1 | -1 | -1 | -1 | -1 | -1 | -1 | -1 | 1 | -1 |
| 114 | 1 | 1 | 1 | -1 | -1 | -1 | 1 | 1 | 1 | 1 | -1 | 1 |
| 115 | 1 | 1 | 1 | -1 | -1 | 1 | -1 | 1 | 1 | 1 | 1 | -1 |
| 116 | 1 | 1 | 1 | -1 | -1 | 1 | 1 | -1 | -1 | -1 | -1 | 1 |
| 117 | 1 | 1 | 1 | -1 | 1 | -1 | -1 | -1 | -1 | 1 | 1 | 1 |
| 118 | 1 | 1 | 1 | -1 | 1 | -1 | 1 | 1 | 1 | -1 | -1 | -1 |
| 119 | 1 | 1 | 1 | -1 | 1 | 1 | -1 | 1 | -1 | 1 | -1 | 1 |
| 120 | 1 | 1 | 1 | -1 | 1 | 1 | 1 | -1 | 1 | -1 | 1 | -1 |
| 121 | 1 | 1 | 1 | 1 | -1 | -1 | -1 | 1 | -1 | 1 | -1 | -1 |
| 122 | 1 | 1 | 1 | 1 | -1 | -1 | 1 | -1 | 1 | -1 | 1 | 1 |
| 123 | 1 | 1 | 1 | 1 | -1 | 1 | -1 | -1 | 1 | 1 | 1 | 1 |
| 124 | 1 | 1 | 1 | 1 | -1 | 1 | 1 | 1 | -1 | -1 | -1 | -1 |
| 125 | 1 | 1 | 1 | 1 | 1 | -1 | -1 | -1 | 1 | 1 | -1 | -1 |
| 126 | 1 | 1 | 1 | 1 | 1 | -1 | 1 | 1 | -1 | -1 | 1 | 1 |
| 127 | 1 | 1 | 1 | 1 | 1 | 1 | -1 | 1 | 1 | -1 | -1 | 1 |
| 128 | 1 | 1 | 1 | 1 | 1 | 1 | 1 | -1 | -1 | 1 | 1 | -1 |
| 129 | 0 | 0 | 0 | 0 | 0 | 0 | 0 | 0 | 0 | 0 | 0 | 0 |
| 130 | 0 | 0 | 1 | 1 | 0 | -1 | 1 | -1 | -1 | 0 | 1 | 1 |
| 131 | 0 | 0 | -1 | -1 | 0 | 1 | -1 | 1 | 1 | 0 | -1 | -1 |
| 132 | 0 | 1 | 0 | 1 | -1 | 1 | 0 | 1 | -1 | 1 | 1 | -1 |
| 133 | 0 | 1 | 1 | -1 | -1 | 0 | 1 | 0 | 1 | 1 | -1 | 0 |
| 134 | 0 | 1 | -1 | 0 | -1 | -1 | -1 | -1 | 0 | 1 | 0 | 1 |
| 135 | 0 | -1 | 0 | -1 | 1 | -1 | 0 | -1 | 1 | -1 | -1 | 1 |
| 136 | 0 | -1 | 1 | 0 | 1 | 1 | 1 | 1 | 0 | -1 | 0 | -1 |
| 137 | 0 | -1 | -1 | 1 | 1 | 0 | -1 | 0 | -1 | -1 | 1 | 0 |

| | | | | | | | | | | | | |
|---|---|---|---|---|---|---|---|---|---|---|---|---|
| 138 | 1 | 0 | 0 | 1 | 1 | 1 | 1 | 0 | 1 | 1 | 0 | 1 |
| 139 | 1 | 0 | 1 | -1 | 1 | 0 | -1 | -1 | 0 | 1 | 1 | -1 |
| 140 | 1 | 0 | -1 | 0 | 1 | -1 | 0 | 1 | -1 | 1 | -1 | 0 |
| 141 | 1 | 1 | 0 | -1 | 0 | -1 | 1 | 1 | 0 | -1 | 1 | 0 |
| 142 | 1 | 1 | 1 | 0 | 0 | 1 | -1 | 0 | -1 | -1 | -1 | 1 |
| 143 | 1 | 1 | -1 | 1 | 0 | 0 | 0 | -1 | 1 | -1 | 0 | -1 |
| 144 | 1 | -1 | 0 | 0 | -1 | 0 | 1 | -1 | -1 | 0 | -1 | -1 |
| 145 | 1 | -1 | 1 | 1 | -1 | -1 | -1 | 1 | 1 | 0 | 0 | 0 |
| 146 | 1 | -1 | -1 | -1 | -1 | 1 | 0 | 0 | 0 | 0 | 1 | 1 |
| 147 | -1 | 0 | 0 | -1 | -1 | -1 | -1 | 0 | -1 | -1 | 0 | -1 |
| 148 | -1 | 0 | 1 | 0 | -1 | 1 | 0 | -1 | 1 | -1 | 1 | 0 |
| 149 | -1 | 0 | -1 | 1 | -1 | 0 | 1 | 1 | 0 | -1 | -1 | 1 |
| 150 | -1 | 1 | 0 | 0 | 1 | 0 | -1 | 1 | 1 | 0 | 1 | 1 |
| 151 | -1 | 1 | 1 | 1 | 1 | -1 | 0 | 0 | 0 | 0 | -1 | -1 |
| 152 | -1 | 1 | -1 | -1 | 1 | 1 | 1 | -1 | -1 | 0 | 0 | 0 |
| 153 | -1 | -1 | 0 | 1 | 0 | 1 | -1 | -1 | 0 | 1 | -1 | 0 |
| 154 | -1 | -1 | 1 | -1 | 0 | 0 | 0 | 1 | -1 | 1 | 0 | 1 |
| 155 | -1 | -1 | -1 | 0 | 0 | -1 | 1 | 0 | 1 | 1 | 1 | -1 |

**Table S2:** $IC_{50}$ analysis of single drug dose response assay compared to mean of other cell lines. Anti-angiogenics in bold.

| **Drug** | **$IC_{50}$ (μM)** | | |
|---|---|---|---|
| | **hSC06** | **hSC31** | **Mean of other cell lines^** |
| Ifosfamide | 8.48 | 19.7 | 10.4* |
| Doxorubicin | 0.000466 | 0.0140 | 0.171 |
| Panobinostat | 0.000107 | 0.00268 | 0.0496 |
| Gemcitabine | 0.0100 | 0.100 | 0.0290 |
| 5-Azacytidine | 0.550 | 43.1 | 9.78* |
| Roybinostat | 0.176 | 14.8 | 4.26* |
| OTX015 | 0.767 | 20.5 | 11.1 |
| Palbociclib | 14.1 | 13.8 | 35.7 |
| **Pazopanib** | 10.9 | 146 | 23.2 |
| Eribulin mesylate | 0.00100 | 0.00549 | 0.0684* |
| Trabectedin | N/D[#] | N/D[#] | 0.00170* |
| Ponatinib | 0.0285 | 0.629 | 1.50 |
| **Sorafenib** | 13.8 | 11.3 | 13.7 |
| **Sunitinib** | 9.88 | 8.69 | 11.6 |
| **Lenvatinib** | 21.8 | 16.4 | 28.5* |
| **Regorafenib** | 25.1 | 13.2 | 9.41* |

**Table S3:** Reported cases of ventricular Solitary Fibrous Tumours/Hemangiopericytomas (SFT/HPCs), to date. We report the 30th known case of ventricular SFT. Notably, with the exception of one other case which developed widespread spinal metastasis and passed on in five months, our case is the only other reported case with recurrence and development of distant metastasis. Hpf = high power field; NA = Not Available/Applicable/ Reported; GTR = Gross Tumour Resection (complete removal of the tumour with negative post-operative margins); STR = Subtotal Tumour Resection (incomplete removal of the tumour with positive post-operative margins).

| Case | Author | Age, Sex | Site of Disease | Type | Size of Disease (Largest Dimension) (cm) | Presentation | Surgical Results | Adjuvant Radiotherapy | Histology (Mitoses, Necrosis, Pleomorphism) | Follow-Up |
|---|---|---|---|---|---|---|---|---|---|---|
| 1 | McDonald and Terry, 1961 | 55, M | Trigone of R lateral ventricle | HPC | 3.5 | L hemiparesis, L quadrantanopia, jargon aphasia | GTR | 50 Gy of focal radiation over 7 days | NA | No evidence of local recurrence or metastasis at 21.5 years |
| 2 | Muttaqin et al., 1991 | 41, M | Trigone of L lateral ventricle | HPC | > 5 | R upper extremity paresthesia, HA, mild speech disturbance | STR | 50 Gy of Local radiation over 30 fractions | High cellularity, staghorn like vascular channels, mitotic index not mentioned | 4 months post-operative readmission for worsening headache |
| 3 | Abrahams et al., 1999 | 40, M | Third ventricle | HPC | 1 | Headache | GTR | Not described | NA | NA |
| 4 | Hattingen et al., 2003 | 43, F | Trigone of L lateral ventricle | HPC | < 3 | Headache | GTR | Not described | Many small vessels, high cell density and plump nuclei without atypia | NA |
| 5 | Al-Brahim et al., 2004 | 53, F | Atrium of R lateral ventricle | HPC | 4.5 | R temporal visual field constriction, L hemiparesis, gait ataxia | GTR | Not described | Mitoses less than 1 hpf, no necrosis, prominent slit-like vasculature with staghorn appearance | NA |
| 6 | Desai et al., 2004 | 40, M | Trigone of R lateral ventricle | HPC | 7 | L homonymous hemianopia, headache | GTR | 60 Gy of local radiation over 30 fractions | Slit-like vascular spaces lined by endothelial cells, no necrosis found in tumour, mitotic activity minimal | No local recurrence or metastasis at 1 year |
| 7 | Bunai et al., 2008 | 9, M | R lateral ventricle | HPC | 0.5 | Headache | Not performed | Not performed | NA | Sudden death thought to be due to neurogenic pulmonary edema caused by the rapid growth of a hemangiopericytoma, with intratumoral hemorrhage |
| 8 | Suzuki et al., 2009 | 31, F | Trigone of R lateral ventricle | HPC | 5 | Left upper quadrantopia, blurred vision, L 6th nerve palsy, headache | GTR | Not performed | Staghorn appearance, mitosis not reported | No local recurrence or metastasis at 4 years |
| 9 | Sumi et al., 2010 | 65, F | Body of bilateral lateral ventricles | HPC | 5 | Gait disturbance | GTR | 55 Gy local radiation | 3-5 mitoses per hpf, absence of necrosis | No local recurrence at 5 years |
| 10 | Tanaka et al., 2011 | 67, F | Trigone of L lateral ventricle | HPC | 6 | R quadrantanopia, R hemiparesis, motor aphasia | GTR | Not performed | Staghorn vessels, no necrosis and mitoses reported | No local recurrence or metastasis at 2 years |
| 11 | Krajewski et al., 2015 | 19, M | R lateral ventricle | HPC | Not Described | L hemiparesis, Headache, nausea | GTR | Fractionated radiation – radiation dose ND | NA | Death 5 months postoperatively with widespread spinal metastasis |
| 12 | Towner et al. 2016 | 23, M | Transcortical parieto-occipital | HPC | 6 | R homonymous hemianopia, word-finding difficulty, R paresthesia, headache | GTR | 60 Gy of local radiation over 30 fractions | 16 mitoses per hpf, multifocal necrosis and anaplasia (WHO Grade III) | No local recurrence or metastasis at 9 months |
| 13 | Yang et al. 2019 | 52, M | Fourth Ventricle | SFT/HPC | 3.3 | Dizziness and progressive weakness in his left extremities | GTR | Routine Radiotherapy (not described in further detail) | Mitotic figures, STAT6 mutation | NA, discharged in good condition on Week 4 post-operatively |
| 14 | Wang et al. 2012 | 52, M | Fourth ventricle, compression of medulla, cerebellum and foramen of Luschka | SFT | 4.5 | Progressive weakness and numbness in all extremities | GTR | | Absent pleomorphism, necrosis and mitosis | No recurrence at one year follow-up |
| 15 | Clarençon et al. 2006 | 32, F | Fourth ventricle | SFT | 2.5 | NA | NA | NA | NA | NA |
| 16 | Cummings et al. 2001 | 52, M | Fourth ventricle | SFT | NA | NA | NA | NA | NA | Autopsy |

| | | | | | | | | | | |
|---|---|---|---|---|---|---|---|---|---|---|
| 17 | Gessi et al. 2006 | 63, F | Fourth ventricle | SFT | 2 | sporadic episodes of vomiting, headaches and vertigo, mild left dysmetria | NA | NA | Absent mitosis and necrosis, pleomorphism not reported, staghorn vessels absent | NA |
| 18 | Kim et al. 2004 | 49, F | Fourth ventricle | SFT | NA | Chronic Headache (months) | GTR | Not described | Mild pleomorphism, rare mitosis, necrosis not reported | No recurrence after one year |
| 19 | Montano et al. 2010 | 61, M | Fourth ventricle | SFT | NA | dizziness, nausea and gait imbalance | GTR | Not described | NA | No recurrence after two years |
| 20 | Sawauchi et al. 2003 | 57, M | Fourth ventricle | SFT | NA | Headache and Nausea | GTR | | NA | NA |
| 21 | Tihan et al. 2003 | Not found, 2 patients | Lateral ventricles | SFT | Not found | Not found | Not found | Not found | NA | Not found |
| 22 | Clarençon et al. 2007 | 44, F | Right Lateral ventricle | SFT | 3.5 | Retro-orbital pain and seizure | NA | NA | NA | NA |
| 23 | Liao et al. 2009 | NA | Right Lateral ventricle | SFT | 5 | NA | NA | NA | NA | NA |
| 24 | Mekni et al. 2009 | 40, M | Right Lateral ventricle | SFT | 5 | NA | NA | NA | Marked pleomorphism, mitosis 5/10 HPF, Focal necrosis | No recurrence after three years |
| 25 | Surendrababu et al. 2006 | 55, F | Left lateral ventricle | SFT | NA | Single episode generalized tonic clonic seizures with post-ictal weakness on the right side (improved on admission) | GTR | NA | Absent pleomorphism, necrosis and mitosis not reported | No recurrence after one year |
| 26 | Vassal et al. 2011 | 60, F | Left lateral ventricle | SFT | 2.5 | 1 month history of gradually progressive speech disturbances and headaches<br><br>Neurological examination at admission revealed Wernicke aphasia with neologisms and moderate weakness of the right side. | GTR | NA | Absent pleomorphism, necrosis and mitosis | No evidence of recurrence after two years |
| 27 | Wright et al. 1993 | 11, F | Right Lateral ventricle | SFT | Not reported | NA | NA | NA | NA | Not reported |
| 28 | Kocak et al. 2004 | 63, M | Third Ventricle | SFT | 2.5 | Weakness of lower extremities and headaches | GTR | NA | NA | No symptoms after 3.5 years |
| 29 | Kinfe et al. 2008 | 75, F | Foramen of Munro | SFT | 2.5 | Occlusive hydrocephalus including cognitive deficits, urine incontinence and gait disturbance | NA | NA | NA | No recurrence after one year, but with moderate residual cognitive deficits |
| 30 | **Our Case, 2022** | 20s, F | Left Lateral Ventricle | HPC | 0.78 | Severe headache | STR | Post-operative radiotherapy (55.8 Gy in 30 fractions) but stopped on the 9th fraction due to poor wound healing; Stereotactic Radiosurgery due to recurrence | First Presentation: WHO Grade II SFT/HPC with staghorn blood vessels, moderate pleomorphism and a mitotic figure of 1 per 10 hpf<br><br>During Recurrence: Grade III SFT/HPC with elevated mitoses at 5 per 10 hpf, invasion of the cerebral cortex by the tumour, very little intervening collagenous stroma between the tumour fascicles, geographic necroses and staghorn vessels | Multiple local recurrences and metastatic disease to bone and liver within 6 years |
